# Leveraging machine learning and self-administered tests to predict COVID-19: An olfactory and gustatory dysfunction assessment through crowd-sourced data in India

**DOI:** 10.1101/2021.10.20.21265247

**Authors:** Ritesh Kumar, Maneet Singh, Prateek Singh, Valentina Parma, Kathrin Ohla, Shannon B Olsson, Varun Saini, Jyotsna Rani, Kunal Kishore, Priyanka Kumari, Parul Ichhpujani, Anupma Sharma, Saurav Kumar, Manu Sharma, Amol P Bhondekar, Anamika Kothari, Viren Sardana, Sudarshan Iyengar, Debasis Dash, Rishemjit Kaur

## Abstract

It has been established that smell and taste loss are frequent symptoms during COVID-19 onset. Most evidence stems from medical exams or self-reports. The latter is particularly confounded by the common confusion of smell and taste. Here, we tested whether practical smelling and tasting with household items can be used to assess smell and taste loss. We conducted an online survey and asked participants to use common household items to perform a smell and taste test. We also acquired generic information on demographics, health issues including COVID-19 diagnosis, and current symptoms. We developed several machine learning models to predict COVID-19 diagnosis. We found that the random forest classifier consistently performed better than other models like support vector machines or logistic regression. The smell and taste perception of self-administered household items were statistically different for COVID-19 positive and negative participants. The most frequently selected items that also discriminated between COVID-19 positive and negative participants were clove, coriander seeds, and coffee for smell and salt, lemon juice, and chillies for taste. Our study shows that the results of smelling and tasting household items can be used to predict COVID-19 illness and highlight the potential of a simple home-test to help identify the infection and prevent the spread.

## 1 INTRODUCTION

Severe acute respiratory syndrome coronavirus 2 (SARS-CoV-2) commonly known as COVID-19, has perhaps been one of the most catastrophic pandemics. In India, the first case was reported in January 2020 [Andrews et al., 2020]. Due to high transmission rates among humans [Ke et al., 2020], it has infected more than 33 million people (according to the latest update available on Sep 8th, 2021) and caused the casualties of more than 0.4 million in India [Worldometers, 2021]. The major challenges that have been posed by COVID-19 are the unspecificity of most symptoms [Gerkin et al., 2021] and the scarcity of facilities permitting mass testing for the SARS-CoV-2 virus [Momtazmanesh et al., 2020, Altayb et al., 2020], which is essential to identify infections and reduce the fatality rate [Rong et al., 2020, Czeisler et al., 2020], especially in developing countries. Hence, there is a need to incorporate other methods to alleviate the load on the existing approaches.

In line with the recent trends in medicine [Sajda, 2006], several studies used machine learning models to predict the diagnosis of COVID-19. These models have been trained using features like images of CT scan and X-ray [Owais et al., 2021, Wang et al., 2021], clinical features [Zoabi et al., 2021, Menni et al., 2020], routine laboratory tests [Feng et al., 2021], demographic as well as symptomatic information [Zoabi et al., 2021] or a combination of these features [Mei et al., 2020]. It has been observed that loss of smell and taste in the majority of COVID-19 infected individuals, appear before other symptoms [Samaranayake et al., 2020]. Lechien et al. [2020a] found that more than 80% out of 417 confirmed COVID-19 patients had reported olfactory dysfunction. Makaronidis et al. [2020] analyzed the number of people who had developed COVID-19 antibodies out of 567 participants from London with a newly developed loss in their sense of smell or taste. Around three fourth (78%) of people with smell and taste loss were found to have COVID-19 antibodies. Out of these, 40% of the people had neither cough nor fever. The participants with the loss of smell were 3 times more likely to have COVID-19 antibodies, compared with those with loss of taste. Working on similar lines, Parma et al. [2020] conducted a global online survey to assess smell, taste, and chemesthesis (often summarized as chemosensory) abilities in participants with COVID-19 and those with other respiratory illnesses. They found that COVID-19 affects not only smell but also taste and chemesthesis. These works have led to the use of various smell and taste tests such as Sniffin Sticks [Lechien et al., 2020b], University of Pennsylvania Smell Identification Test [Moein et al., 2020], Brief Smell Identification Test, Waterless Empirical Taste Test for the objective evaluation of chemosensory dysfunction and provide insights into the characteristics of the disease [Cao et al., 2021]. The main advantage of these tests is their portability and scalability. However, they require close physical contact between the patient and medical assistant as well as the test material (e.g. the Sniffin sticks), which both pose high risks of spreading the virus further. Hence, alternative testing methods that permit social distancing are urgently needed.

In this work, we have presented a self-administered smell and taste test with common household items such as spices, tea, and dairy products to test its suitability to predict COVID-19 illness. Using smell and taste along with other health-related features, we compared model performances and identify the most informative features. The use of machine learning models to detect COVID-19 based on the earliest symptoms is necessary for developing countries like India where not all citizens have immediate access to medical care or testing facilities.

Thus, our study sought to answer the following research questions:-

- ***RQ1: Can we design a machine learning model based on various generic features including demographic (age and gender), smoking habit, number of daily physical contacts, health issues (obesity, diabetes, etc), along with general symptoms such as fever, headache, etc. and smell and taste specific symptoms (complete loss of smell, change in taste of bitter, etc) for efficient prediction of COVID-19 diagnosis?***
- ***RQ2: Can self-administered tests performed at home be used to predict COVID-19 and which of the item groups of the self-administered tests are significantly effective in diagnosing the patient as being COVID-19 positive or negative?***

From our study, we found that the random forest model trained on self-administered olfactory and gustatory test ratings along with the generic features performed better than the models trained on the various generic features (i.e. demographic, health issues, symptoms, etc.). The ratings obtained from olfactory and gustatory based self-administered tests for household items like salt, chilli, etc. (for taste) as well as spices, coffee, etc. (for smell) were found to be significantly different for individuals diagnosed as COVID-19 positive than that of the individuals who reported themselves as negative. The novelty of this paper is to show that machine learning models built upon the self-reports and the objective taste and smell based ratings using household items could be used to predict COVID-19. This could be useful for a resource-constrained country like India where models can act as assistive diagnosis methods for rapid assessment.

## 2 EXPERIMENTAL DESIGN

The overall design of the study for answering our key research questions is shown in Figure 1. First, the data from the participants was collected by organizing an online survey through a web application. The survey was approved by the institutional ethics committee (IEC/CSIO/2020 No. 23 dated June 18, 2020 and No. 30 dated September 23, 2020). The survey included an informed consent form and only the participants aged above 18 years were allowed. The data included for this study was from 25th Aug 2020 to 9th May 2021. To answer RQ1 stated above, we first extracted the statistically significant features from demographics, smoking habits, number of daily physical contacts, health issues, general symptoms and smell and taste specific symptoms (coded as GEN for statistically significant Generic features). These features were then used to develop and assess multiple machine learning models for the prediction of disease. Similarly to answer RQ2, we first identified various olfactory and gustatory groups, that are significantly effective in COVID-19 diagnosis, through the statistical analysis on the ratings obtained from self-administered tests for COVID-19 positive and negative individuals (coded as OBSAT for olfactory based self-administered test results and GBSAT for gustatory based self-administered test results). Later different combinations of these features were used to train different machine learning models and a comparison was done to assess the efficacy of these features for the diagnosis of COVID-19.

**Figure 1.**
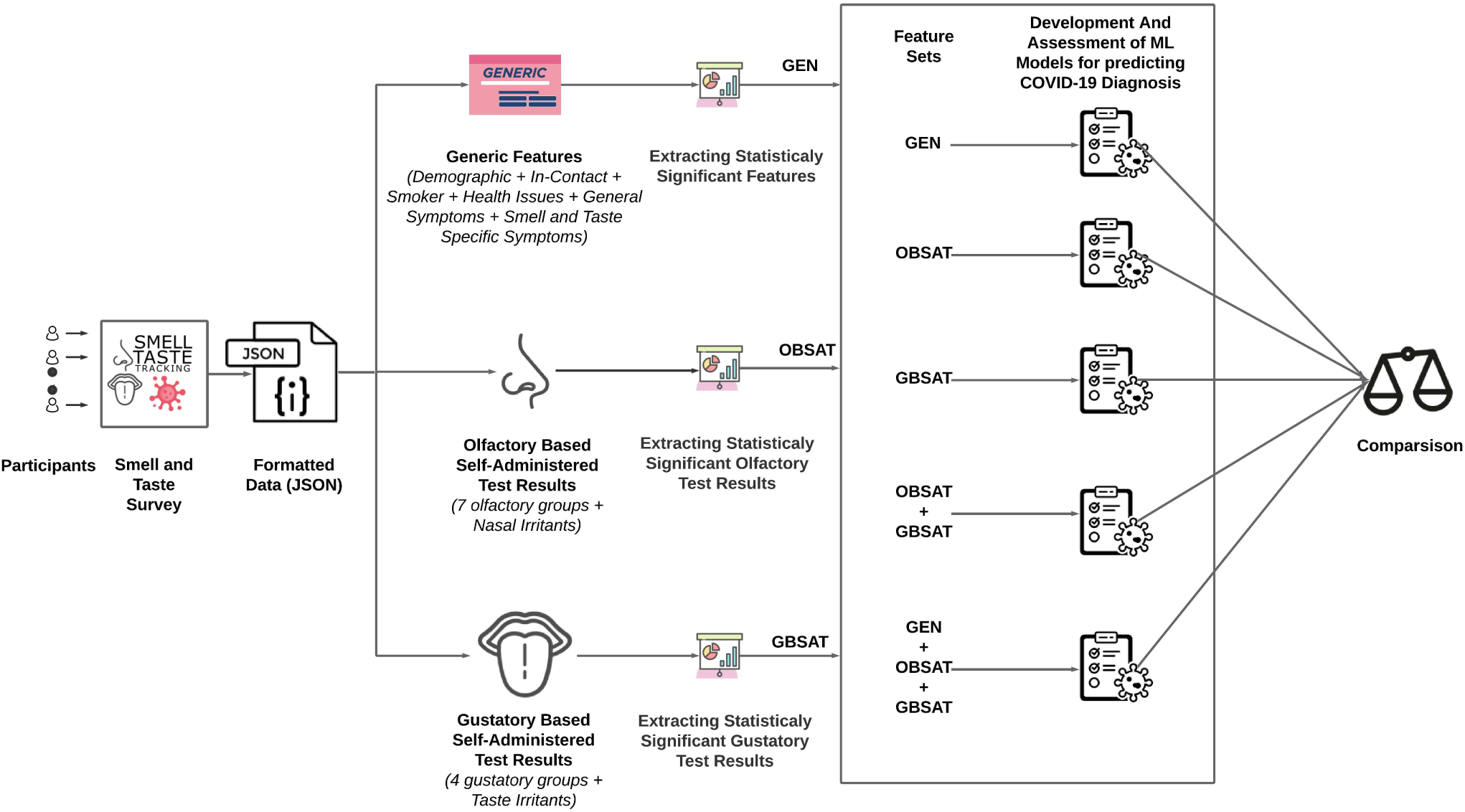
The overall workflow of our study. The significant generic features (GEN) were extracted and used in prediction task for answering RQ1. Similarly, to answer RQ2, first the significant olfactory (OBSAT) and gustatory (GBSAT) based self-administered test results were extracted. Lastly, different combinations of these features were used for comparing their prediction capability in diagnosis of COVID-19. Here ML stands for Machine Learning.

## 3 MATERIALS AND METHODS

In this section, we have discussed various methods employed in our study including data collection as well as development and evaluation of the machine learning models for predicting the diagnosis of COVID-19.

### 3.1 Data

We built a smell and taste tracking web app using open-source software vue.js, Flask, Python and JavaScript. The web app was deployed at https://mapcorona.in/. In the web app, the participants were asked several questions pertaining to their demographic (i.e., year of birth, gender and country of residence), the number of people they came in contact with, their smoking habit, health issues (in the past six months), information related to various symptoms (general or smell and taste specific) in the past fourteen days, their COVID-19 diagnosis and ratings for self-administered olfactory and gustatory tests. The detailed questionnaire is available in the supplementary material.

For self-administered tests, we used a visual analogue scale in the range from 0 to 100 for olfactory and gustatory ratings using household items. In this scale, 0 denotes “No Sensation” and 100 corresponds to “Very Intense”. We provided seven different fragrant items for smelling (with default selection as detergents, clove, coriander, garam masala, lemon, coffee and milk primarily because of the ease of availability) and 4 different edible items for tasting (with salt, sugar, lemon and tea as default selection) and asked the participants to rate the intensity of their smell and taste respectively.We also selected Vicks VapoRub topical ointment (Procter & Gamble, a topical cough suppressant used in India) which has eucalyptus oil, menthol, camphor as major constituents to observe olfactory irritant sensations along with chilli for taste irritants. Table 1 shows the olfactory groups and the items selected for the test. The primary reason for the selection of items for odour tests was the presence of molecules that activated major groups of receptors along with ease of availability, e.g. clove, nutmeg which has primarily eugenol; it triggers as many as 45 olfactory receptors [Horio et al., 2019]. One of the groups consisting of soaps, detergents have aldehydes and some other sulphur related compounds which trigger a wide range of olfactory receptors [Bak et al., 2019]. The spice mixture related group was chosen to cater to the broad sensory activation by various major constituent molecules. Bak et al. [2019] identified 6 major groups of phytochemicals co-activating the known olfactory receptors, our selection of items encompassed the above-mentioned groups. Similarly, Table 2 specifies the set of items used for different taste groups.

**Table 1.**
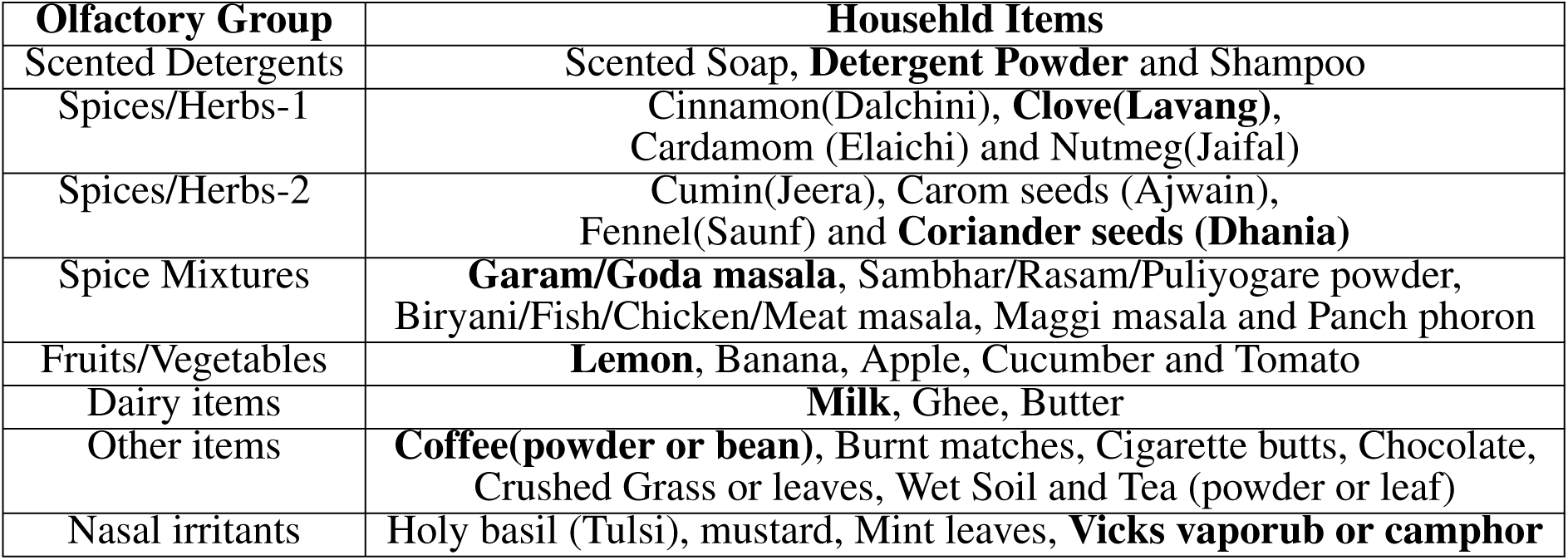
Household items for self-administered test for olfactory assessment. Items provided as default selection in each category is marked as bold.

| **Olfactory Group** | **Househld Items** |
| --- | --- |
| Scented Detergents | Scented Soap, **Detergent Powder** and Shampoo |
| Spices/Herbs-1 | Cinnamon(Dalchini), **Clove(Lavang)**, Cardamom (Elaichi) and Nutmeg(Jaifal) |
| Spices/Herbs-2 | Cumin(Jeera), Carom seeds (Ajwain), Fennel(Saunf) and **Coriander seeds (Dhania)** |
| Spice Mixtures | **Garam/Goda masala**, Sambhar/Rasam/Puliyogare powder, Biryani/Fish/Chicken/Meat masala, Maggi masala and Panch phoron |
| Fruits/Vegetables | **Lemon**, Banana, Apple, Cucumber and Tomato |
| Dairy items | **Milk**, Ghee, Butter |
| Other items | **Coffee(powder or bean)**, Burnt matches, Cigarette butts, Chocolate, Crushed Grass or leaves, Wet Soil and Tea (powder or leaf) |
| Nasal irritants | Holy basil (Tulsi), mustard, Mint leaves, **Vicks vaporub or camphor** |

**Table 2.** Household items for self-administered test for gustatory assessment. Items provided as default selection in each category is marked as bold.

| **Gustatory Group** | **Household Items** |
| --- | --- |
| Sweet | **Sugar** and Sweetener |
| Salty | **Salt** |
| Sour | **Lemon**, Tamarind, Kokam, Vinegar (any kind but balsmic) |
| Bitter | **Tea (leaves or grains or dust)**, Neem leaves, Coffee (beans, grounds, instant powder) and Fenugreek |
| Taste irritants (cooling, tickling, burning or stinging sensation) | **Chilli**, Black pepper, ENO (fruit salt), Mint leaves |

The questionnaire was distributed through social media and word of mouth. Every participant tasted or smelt different household items following a protocol and accordingly rated the perceived intensity of the sensation on the Visual Analog Scale (Please refer to Questions No 19 to 31 in the supplementary material). Overall, we collected data from 249 Indian participants. We further used the following exclusion criteria:- (1) participants having pending COVID-19 test report (No. of participants = 2) and/or (2) the ones with inconsistent reporting regarding smell or taste (No. of participants = 5) and/or (3) those suffering or have suffered from a prior smell or taste disorder (Please refer to Question No. 14 in the supplementary material) in the past six months (No. of participants = 3). The resultant data consisted of 239 participants (Male = 163, Female = 76, age: *μ* = 36.73, *σ*= 12.60). Out of those 239, 105 participants were diagnosed with COVID-19 and the rest reported themselves as negative. As some of the questions in the survey were left unanswered by many of the respondents, therefore, we filtered them and used the remaining features (listed in Figure 2) with missing values filled with median [Acuna and Rodriguez, 2004].

**Figure 2.**
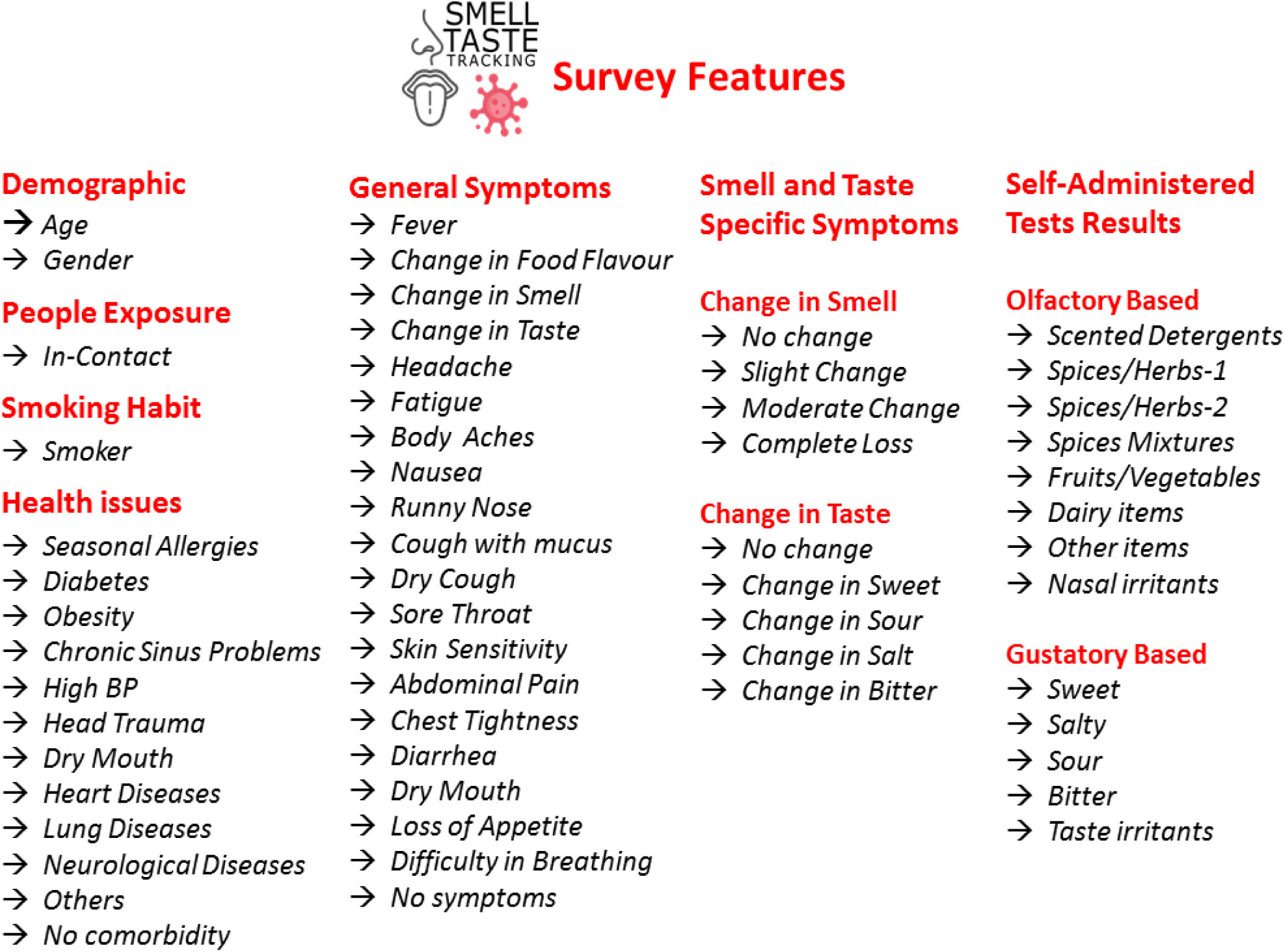
Categorization of several features of the participants obtained from the survey.

### 3.2 Model Development and Assessment

Here, we have described the process of developing and evaluating the models. The implementation was done using the python *sklearn* library. It mainly involves the following two phases:-

#### 3.2.1 Data Preprocessing

As shown in Figure 2, our data consisted of 59 features. Out of all these features, age and the self-administered test ratings were numerical and the remaining were categorical. The numerical features were transformed using standardization, whereas, each categorical feature was converted into multiple binary features using a one-hot encoding method. One hot encoding transforms a single variable with n observations and d distinct values, to d binary features with n observations each [Lantz, 2019]. Thus, on applying one-hot encoding, there were a total of 69 features (including both binary as well as numerical) for each participant.

#### 3.2.2 Model Selection

In this phase, we have compared the performance of five machine learning algorithms namely Naïive Bayes, Decision Trees, Random Forest, Logistic Regression and Support Vector Machines to predict COVID-19 diagnosis. The optimal values of the parameters of these algorithms were obtained using the grid search method with F1-score as the maximising criteria (Please refer to Table S1 given in the supplementary material). The performance of a machine learning model is generally assessed using a point estimate of the evaluation metric such as accuracy or F1-Score. Here, a single value might not be a good indicator of the prediction capability, hence multiple sample sets from the given data are generated to train and test the model for capturing variations in the performance of the model. The percentile bootstrap method [Efron and Tibshirani, 1994] was employed in our study for obtaining confidence intervals as performance estimates (Refer Figure S1 in Supplementary Material Part A). It is a non-parametric method used for computing standard error and finding a confidence interval for a given statistic. In our case, the statistic would be the evaluation metrics used for a machine learning model, i.e. Accuracy, Recall, Precision and F1-Score. For doing so, 1000 bootstraps, i.e. training and test sets, were generated using sampling with replacement approach. Thus the given model was executed for all the bootstraps and the lower and upper bound estimates (using alpha=0.05, i.e. 95% confidence interval) were obtained for all the evaluation metrics.

## 4 RESULTS AND DISCUSSIONS

In this section, we have reported and discussed the findings related to two key research questions of our study.

### 4.1 Can we design a machine learning model based on various generic features including demographic (age and gender), smoking habit, number of daily physical contacts, health-related (obesity, diabetes, etc), along with general (fever, headache, etc) and smell and taste specific (complete loss of smell, change in taste of bitter, etc) symptoms for efficient prediction of COVID-19 diagnosis?

In order to answer the research question, we first aimed at identifying the generic features that are significantly capable (*α* = 0.05) of differentiating infected individuals from non-infected ones. The category-wise results of statistical analysis are given as follows:-

- **Demographic:** The comparison between COVID-19 and Non-COVID-19 participants on the basis of demographic features i.e. gender and age can be seen in Figures S2A and S2D (Supplementary Material Part A) respectively. The statistical tests were also conducted on both features. For gender, Pearson’s chi-square test [Pearson, 1900] was done, whereas, in the case of age, we used Mann Whitney U test [Mann and Whitney, 1947] as age is numeric in nature with non-gaussian distribution (Figure S2D). The gender did not play a significant role (*χ*^2^ statistic = 0.3490 and p-value = 0.5547) in the determination of the COVID-19 and Non-COVID19 infected patients. This has been observed in some other studies as well [Mihaltan et al., 2021]. On the other hand, the younger population were comparatively lesser in proportion among the infected participants (*ρ* statistic = 5395.5 and p-value = 0.0001).
- **People Exposure:** Here, we have compared the number of persons with whom an individual comes in contact i.e. *In-Contact* (Figure S2B in Supplementary Material Part A). Interestingly, based on the statistical analysis, the *In-Contact* feature seems to be one of the essential factors that distinguish a COVID-19 diagnosed person from others (*ρ* statistic = 14.869 and p-value = 0.0109). This is in consonance with some of the previous works where avoidance of crowds has been shown to be an effective measure to control the spread of infectious disease [Lau et al., 2003].
- **Smoking Habit:** The habit of smoking (*Smoker*) among the COVID-19 and Non-COVID-19 participants was also compared (Figure S2C in Supplementary Material Part A). The results of statistical analysis, did not find the *Smoker* feature to play any significant role in the diagnosis of COVID-19 (*χ*^2^ statistic = 1.419 and p-value = 0.7011).
- **Health Issues:** We compared the COVID-19 and Non-COVID-19 participants based on the health issues reported by them in the survey for the past six months. As seen from Figure S3 (Supplementary Material Part A), there were no COVID-19 positive individuals who suffered from any lung disease or diseases other than the ones mentioned in our data. Similarly, there were no COVID-19 negative participants who suffered from any heart disease. None of the health issues was dominant, as found after performing chi-square tests of significance involving both types of participants (Table 3). However, the number of participants who reported to have been diagnosed with COVID-19 and co-morbidity was more than that of the non-COVID-19 participants (Table 3). The previous studies have associated the existence of health issues with the risk of getting severe COVID-19 [Wang et al., 2020].

**Table 3.** Statistical results for comparing COVID-19 and non-COVID-19 participants with respect to health issues observed in the last six months. Here, the significant results are highlighted in bold.
- **General Symptoms:** The symptomatic differences in the participants can be seen in Figure S4 (Supplementary Material Part A). Here, symptoms like fever, change in food flavour, change in smell, change in taste, cough with mucus, chest tightness and breathing difficulty were found to be present significantly more in the case of participants that were infected with the virus (Table 4). None of the symptoms was significantly higher in Non-COVID-19 participants. In our survey data, the number of infected individuals who reported not having any symptoms were relatively lesser (around 10%) as compared to their counterpart (around 40%) as observed in Figure S4. This indicates that even the presence of any one of the symptoms is an indication that an individual should get tested for COVID-19.

| Health Issues (last 6 months) | $\chi^2$ Statistic | p-value |
| --- | --- | --- |
| Seasonal Allergies | 1.2613 | 0.2614 |
| Diabetes | 0.0048 | 0.9446 |
| Obesity | 1.1184 | 0.2902 |
| Chronic Sinus Problems | 1.4875 | 0.2352 |
| High BP | 0.0054 | 0.9413 |
| Head Trauma | 0.0454 | 0.8313 |
| Dry Mouth | 0.0048 | 0.9446 |
| Heart Diseases | 0.0150 | 0.9025 |
| Lung Diseases | 0.0000 | 1.0000 |
| Neurological Diseases | 0.0000 | 1.0000 |
| Other | 0.9170 | 0.3383 |
| No Co-morbidity | **21.2420** | **0.0000** |

**Table 4.** Statistical results for comparing COVID-19 and non-COVID-19 participants with respect to symptoms observed in the last fourteen days. Here, the significant results are highlighted in bold.
- **Smell and Taste Specific Symptoms:** We assessed the reported smell and taste change on 4 scales (no change, slight change, moderate change and complete loss for smell change) and basic taste features (Sour, Bitter, Salty, Sweet and no change). As shown in Figure S5 (Supplementary Material Part A), both the symptoms highlighted differences between COVID-19 and Non-COVID-19 participants. Further, we performed statistical tests to ascertain if the reported changes were significant, it was observed that the majority of the COVID-19 infected participants reported partial or complete loss of sense of smell (Table 5). On the other hand, the changes in taste of sweet, salt, bitter and sour were also more in the case of COVID-19 participants. This further strengthens the results reported earlier regarding the loss of sense of smell and taste being one of the most important symptoms in the diagnosis of the COVID-19 disease [Ceron et al., 2020, Parma et al., 2020]. It also strengthens our case to delve deeper into identifying which of the home items are better indicators in this case.

| General Symptoms | $\chi^2$ Statistic | p-value |
| --- | --- | --- |
| Fever | **65.6068** | **0.0000** |
| Change in Food Flavour | **4.6794** | **0.0305** |
| Change in Smell | **9.3526** | **0.0022** |
| Change in Taste | **11.3862** | **0.0007** |
| Headache | 2.8650 | 0.0905 |
| Fatigue | 0.0305 | 0.8613 |
| Body Aches | 0.0000 | 1.0000 |
| Nausea | 0.0000 | 1.0000 |
| Runny Nose | 3.6815 | 0.0550 |
| Cough with mucus | **29.0047** | **0.0000** |
| Dry Cough | 1.1360 | 0.2865 |
| Sore throat | 0.6842 | 0.4081 |
| Skin Sensitivity | 2.3873 | 0.1223 |
| Abdominal Pain | 0.5405 | 0.4622 |
| Chest Tightness | **13.1800** | **0.0003** |
| Diarrhea | 0.0000 | 1.0000 |
| Dry Mouth | 0.0000 | 1.0000 |
| Loss of Appetite | 0.0048 | 0.9446 |
| Difficulty in Breathing | **8.4278** | **0.0036** |
| No Symptoms | **23.9219** | **0.0000** |

**Table 5.**
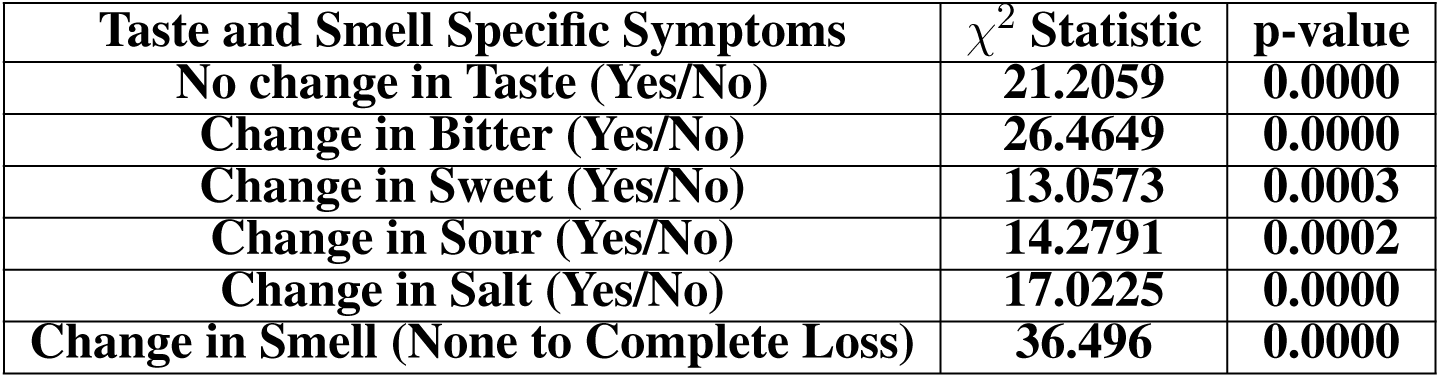
Statistical comparison of smell and taste specific symptoms for COVID-19 and Non-COVID-19 participants. Here bold entries corresponds to significant results.

| Taste and Smell Specific Symptoms | $\chi^2$ Statistic | p-value |
| --- | --- | --- |
| No change in Taste (Yes/No) | **21.2059** | **0.0000** |
| Change in Bitter (Yes/No) | **26.4649** | **0.0000** |
| Change in Sweet (Yes/No) | **13.0573** | **0.0003** |
| Change in Sour (Yes/No) | **14.2791** | **0.0002** |
| Change in Salt (Yes/No) | **17.0225** | **0.0000** |
| Change in Smell (None to Complete Loss) | **36.496** | **0.0000** |

We further set out to model and evaluate machine learning algorithms using a total of 25 statistically significant generic features for the prediction task. In this regard, five machine learning models (i.e. Naïive Bayes, Decision Trees, Random Forest, Logistic Regression and Support Vector Machines) were trained and assessed using the method discussed in the previous section. The performance of all the models on the significant generic features of the survey participants is compared (Table 6). Here we have given lower bounds as well as the upper bound score for each evaluation metric, with the confidence interval of 95 %. Out of the different machine learning classifiers, the random forest model seems to perform comparatively better than the other models. The lower bound score is higher for every evaluation metric in the case of the random forest model. Even for the upper bound score, the difference between the performance of the random forest model and the model with the maximum score is minimal. Another important thing to note here is the difference in the lower and upper bound values is also around 10% for random forest classifiers. This ensures that the model is highly stable i.e. lesser variations in the prediction performance. The scores (lower bound and upper bound) for all four evaluation metrics for the logistic regression model as well as support vector machines are close to the random forest model.

**Table 6.**
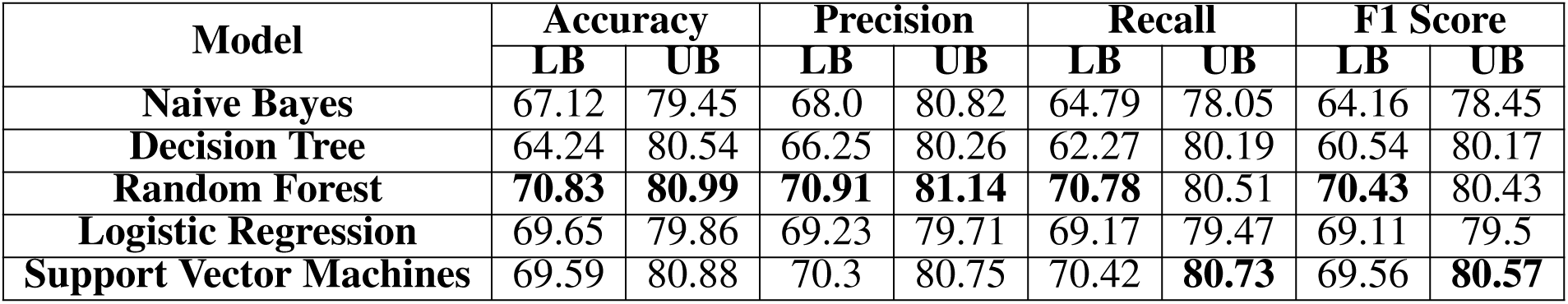
Performance indices with respect to each of the five machine learning models trained on significant generic features (LB-Lower Bound and UB-Upper Bound). The scores marked with bold indicates the maximum value achieved for lower bound or upper bound out of five models.

| Model | Accuracy |  | Precision |  | Recall |  | F1 Score |  |
| --- | --- | --- | --- | --- | --- | --- | --- | --- |
|  | LB | UB | LB | UB | LB | UB | LB | UB |
| Naive Bayes | 67.12 | 79.45 | 68.0 | 80.82 | 64.79 | 78.05 | 64.16 | 78.45 |
| Decision Tree | 64.24 | 80.54 | 66.25 | 80.26 | 62.27 | 80.19 | 60.54 | 80.17 |
| Random Forest | **70.83** | **80.99** | **70.91** | **81.14** | **70.78** | 80.51 | **70.43** | 80.43 |
| Logistic Regression | 69.65 | 79.86 | 69.23 | 79.71 | 69.17 | 79.47 | 69.11 | 79.5 |
| Support Vector Machines | 69.59 | 80.88 | 70.3 | 80.75 | 70.42 | **80.73** | 69.56 | **80.57** |

### 4.2 Can self-administered tests performed at home be used independently to predict COVID-19 and which of the item groups of the self-administered tests are significantly effective in diagnosing the patient as being COVID-19 positive or negative?

Out of the several household items used in our study, we are interested in identifying those items that are statistically able to differentiate between COVID-19 positive and negative individuals. In this regard, the results obtained from the olfactory and gustatory based self-administered tests for all the survey participants were used to assess their role in differentiating between COVID-19 positive and negative individuals. As the ratings for a given olfactory or gustatory product is in the range from 0 to 100, therefore we compared the estimated density distribution of both COVID-19 and Non-COVID-19 participants (Figure 3 and Figure 4). It can be seen that participants suffering from COVID-19 have a higher probability of experiencing a low sense of smell as well as taste as compared to the Non-COVID-19 participants. Similarly, ones not infected with the virus mainly observed higher intensity ratings of smell and taste. We also conducted a statistical test (*α* = 0.05) for each of the features for finding significant differences between both categories of participants. Since none of the plots (in Figure 3 and Figure 4) follows Gaussian distribution, hence Mann Whitney U test was performed. The results of significant analysis for olfactory based tests are given in Table 7. In the case of smell, items such as spices and their mixtures were found to have significantly different test results or intensities for COVID-19 and Non-COVID-19 participants. In the case of gustatory tests, salty, sour and taste irritant items resulted in different test results for COVID-19 and non-COVID-19 patients (Table 8). Recall that for each category of household items, multiple items were included (Table 1 and Table 2). Out of all the items in each category, one which was set as the default selection was the one used by the majority of the participants. Thus items like detergent powder, clove, garam masala, coriander seeds, lemon, milk and coffee were used for smell by the majority of participants. On the other hand, sugar, salt, lemon and tea were the key items used for taste by the participants.

**Table 7.** Statistical comparison of olfactory ratings for participants with and without COVID-19. Here bold entries corresponds to significant results.

| Olfactory Based Groups | $\rho$ Statistic | p-value |
| --- | --- | --- |
| Scented Detergents | 6281.0 | 0.0769 |
| Spices/Herbs-1 | **5608.0** | **0.0034** |
| Spices/Herbs-2 | **5903.0** | **0.0162** |
| Spices Mixtures | **6005.5** | **0.0257** |
| Fruits/Vegetables | 6699.5 | 0.2631 |
| Dairy Items | 6380.5 | 0.1081 |
| Other items | **6008.5** | **0.026** |
| Nasal Irritants | **5128.0** | **0.0001** |

**Table 8.**
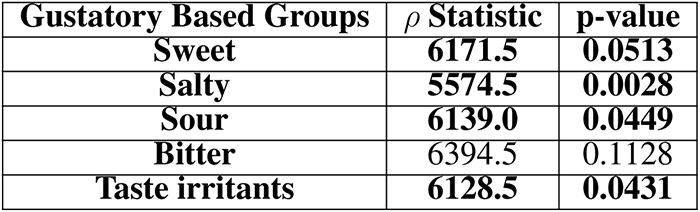
Statistical comparison of gustatory ratings for participants with and without COVID-19. Here bold entries corresponds to significant results.

| Gustatory Based Groups | $\rho$ Statistic | p-value |
| --- | --- | --- |
| Sweet | **6171.5** | **0.0513** |
| Salty | **5574.5** | **0.0028** |
| Sour | **6139.0** | **0.0449** |
| Bitter | 6394.5 | 0.1128 |
| Taste irritants | **6128.5** | **0.0431** |

**Figure 3.**
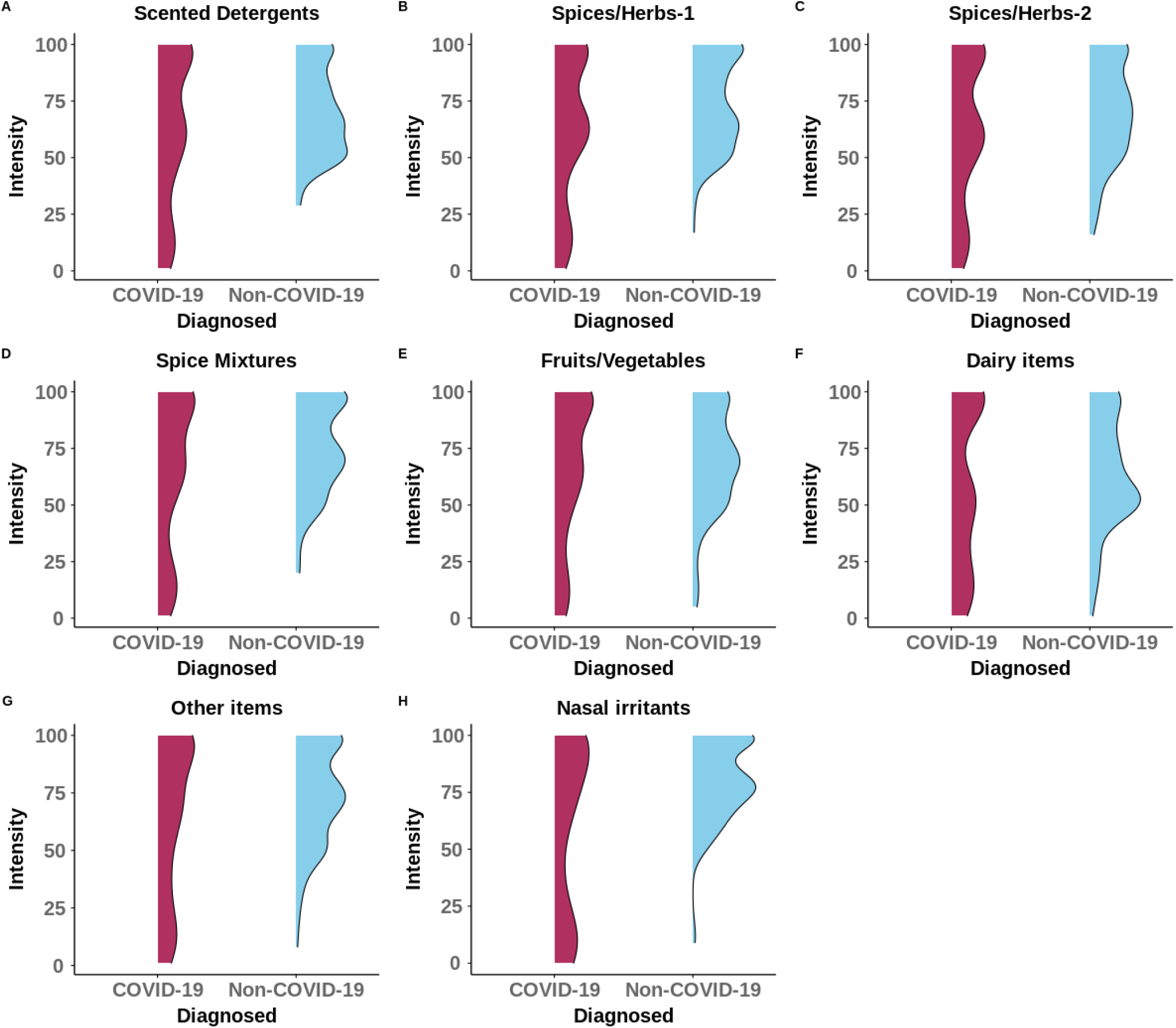
Probability distribution of ratings obtained from self administered tests using household items from gustatory based groups.

**Figure 4.**
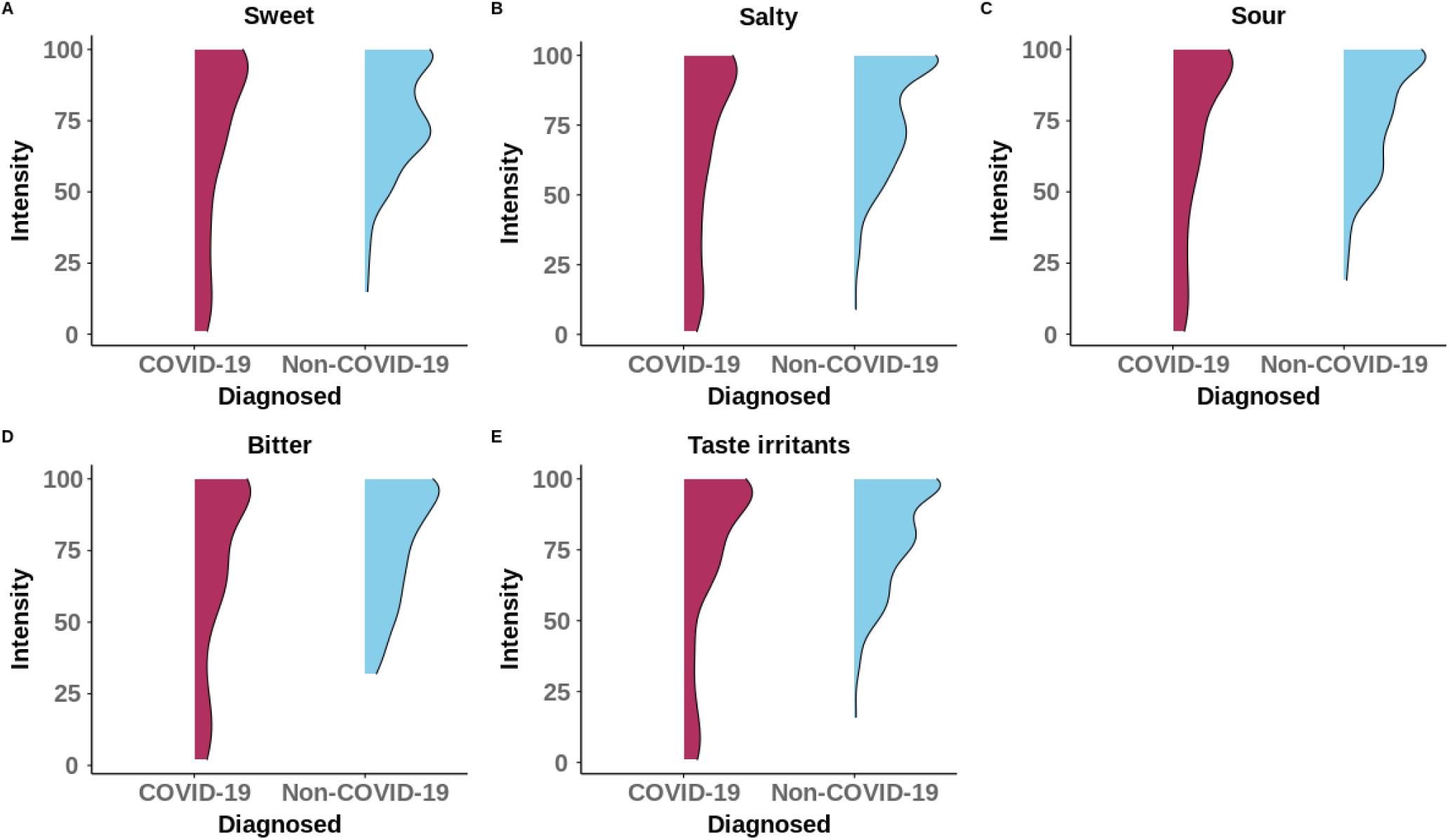
Probability distribution of ratings obtained from self administered tests using household items from gustatory based groups.

These significantly differentiating household items were then used to design ML models for the diagnosis of COVID-19 by generating different subsets of the features. Apart from the generic features (GEN) used previously, we used four other feature sets. It includes olfactory based self-administered test (OBSAT), gustatory based self-administered test (GBSAT), both olfactory and gustatory based self-administered test (OBSAT+GBSAT) and a combination of all the features (GEN+OBSAT+GBSAT). Similar to generic features, all these four feature sets were also used independently for the development and assessment of machine learning models for the prediction of COVID-19. The results of the prediction of these four sets are presented in Table 9. Overall, the random forest classifier is the best performing machine learning algorithm, amongst all five algorithms, for the detection of the disease. The same was observed previously in the case of models trained on only generic features (Table 6). We have also compared the prediction capability of the overall best performing model (Random forest in our case) on all five feature sets (Figure 5) used in our study. It was found that the model trained on the self-administered test results in combination with generic features of survey participants outperformed all the other models trained on different feature combinations. Thus, olfactory and gustatory based self-administered tests can help in the more efficient prediction of the diagnosis for COVID-19. If we look at the performances of the models trained independently on olfactory and gustatory based tests ratings, the former surpassed the latter for all the evaluation matrices. On combining the ratings of both self-administered tests results, the model prediction performance improved. Thus, both the tests have their importance in the diagnosis of COVID-19. The item groups which have come out to be significantly differentiating between participants who reported having COVID-19 and the ones not having it, throw some light onto the cultural and biological aspect of the olfaction. First, the use of spices is one of the most significant parts of the Indian culture in general and the people are ‘trained’ because of the culinary practices to differentiate spices. Second, since spices are a significant part of the culinary practice and most of the default items as described previously(coriander seeds, garam masala, clove) affect a broad range of receptors, people reporting these to have been affected seems reasonable. While the non-significance of fruits/vegetables and dairy items for differentiating does not seem clear from a biological point of view and needs a deeper understanding and further research.

**Table 9.**
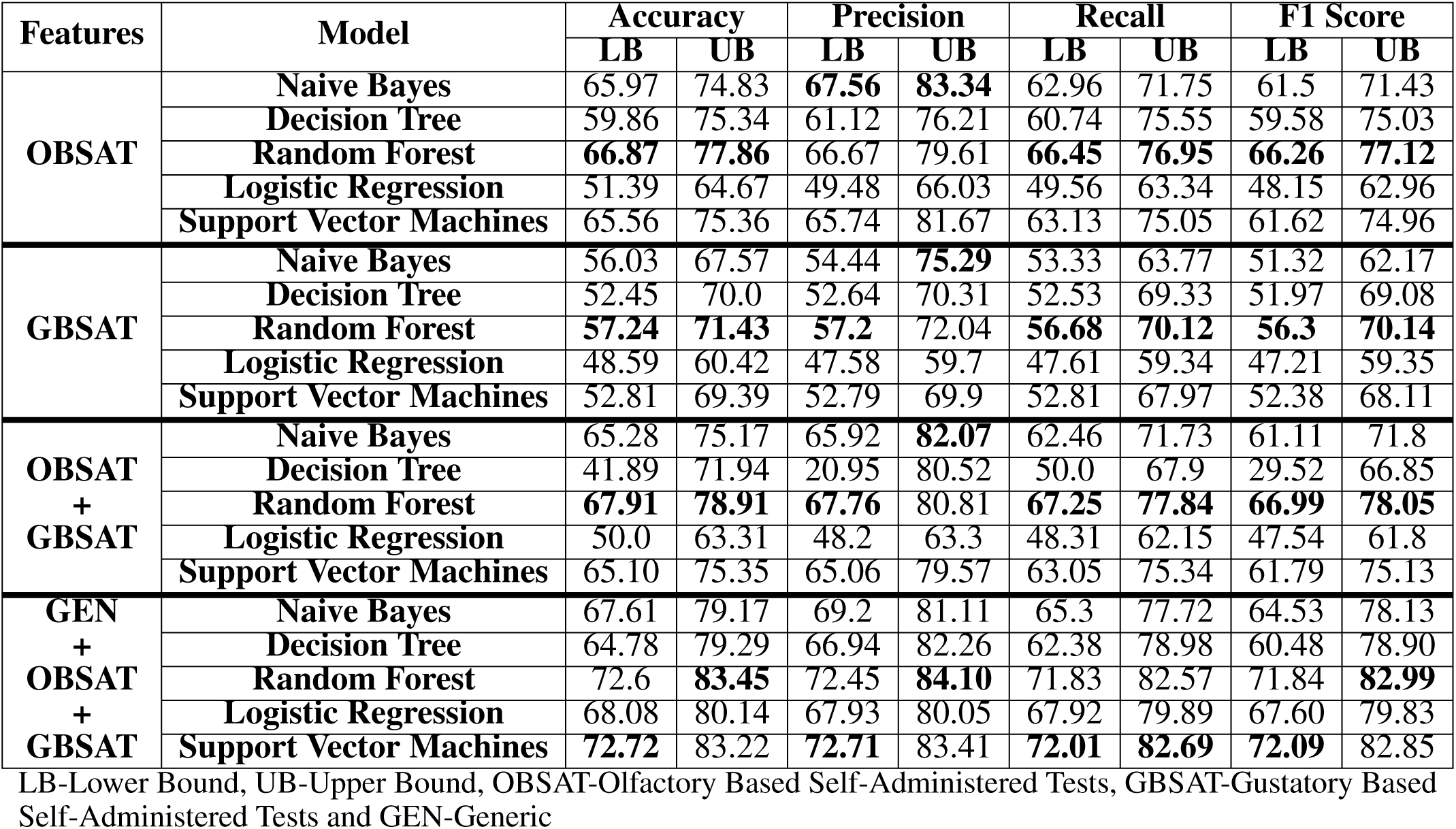
Performance indices for the five models trained on different feature sets.

**Figure 5.**
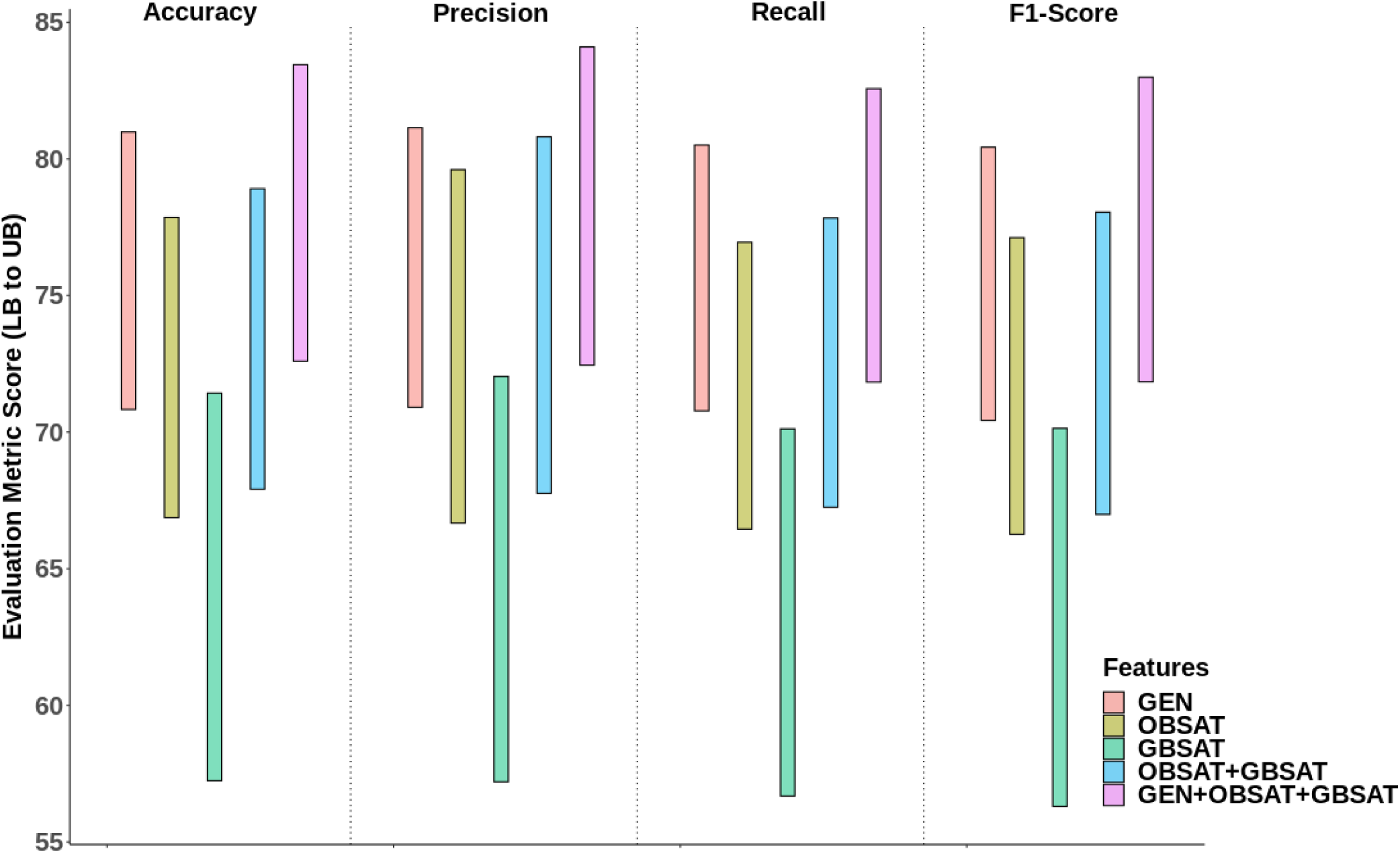
Performance of five feature sets in predicting COVID-19. (generic (GEN), olfactory based self-administered test (OBSAT), gustatory based self-administered test (GBSAT), both olfactory and gustatory based self-administered test (OBSAT+GBSAT) and a combination of all the features (GEN+OBSAT+GBSAT). The lower end of the bar denotes Lower Bound (LB) for each evaluation metric and similarly the upper end of the bar denotes upper bound (UB).

## 5 CONCLUSION

In this work, we conducted an online survey wherein, along with generic information, we conducted olfactory and gustatory based self-administered tests using a variety of household items such as spices, tea, coffee, lemon and milk. Some of the household items such as clove, garam masala and coffee for smell and similarly salt, lemon and tea for taste resulted in statistically significant results for differentiating COVID-19 and Non-COVID-19 participants. Further, self-administered tests were shown to improve the ability to differentiate between positive and negative participants leveraging machine learning algorithms. This hints at the use of customised sniffing sticks or such olfactory tests for different cultures. Our findings may open up this field to conduct more such studies and also incorporate data from different nations to evaluate the importance of self-administered tests at a global scale. All said the online crowdsourcing based survey come with their challenges such as traceability and anonymity. One more limitation of this study is the lack of details regarding the quantum of olfactory loss with the variants of the virus. We believe this research is a window to the stronger case for customised olfactory tests.

## Data Availability

All data and code produced will be available online at
https://github.com/riteshcanfly/homeTestSmellTaste-analysis

## ACKNOWLEDGEMENT

Ritesh Kumar would like to thank the Royal Society, UK for conducting part of this research. It was partly supported by Newton International Fellowship grant NIF *\* R1 *\* 181328. The authors would also like to thank CSIR for facilitating and funding this research. The authors would also like to thank GCCR.

## DATA AVAILABILITY STATEMENT

The anonymised data and code are available on github at Code and figure repo.

