## Supplementary Material B for "Leveraging machine learning and self-administered tests to predict COVID-19: An olfactory and gustatory dysfunction assessment through crowd-sourced data in India"

### SUPPLEMENTARY MATERIAL (PART B)

#### Self-Assessment Test of Smell and Taste

Welcome to taste and smell tracking website. Following a wave of reports about rapid onset of smell loss, health organizations throughout the world have recognized loss of smell and taste as a marker for the COVID-19, even in the absence of other symptoms. Smell and taste loss have been added to the list of biomarkers for early detection of COVID-19 recently by WHO, ICMR-India, CDC, USA and many such health institutes. [1,2,3,4,5,6]

We are conducting a smell and taste survey in which we ask you to smell and taste some readily available home items and report the sensitivity on the website. Through this web app we also strongly encourage you to report any changes in sense of smell and taste along with any future COVID-19 testing and results. It will be help us in determining that if smell and taste sensitivity can identify asymptomatic carriers. We will also be able to determine a cut-off for smell and taste sensitivity that indicates likely infection.

**Reference links**


1. GCCR (2020) More than just smell - COVID-19 is associated with severe impairment of smell, taste, and chemesthesis.
2. Coincidence of COVID-19 epidemic and olfactory dysfunction outbreak.
3. Isolated sudden onset anosmia in COVID-19 infection. A novel syndrome?.
4. Olfactory and Taste Disorders in Patients With Severe Acute Respiratory Coronavirus 2 Infection: A Cross-sectional Study.
5. ICMR expands coronavirus symptoms list; includes loss of taste and smell, muscle pain, diarrhoea.
6. New loss of taste or smell.

### Frequently Asked Questions??

**What is this test?**

In this test we ask you to smell and taste some house-hold items over a period of next three months preferably every week. We are conducting this survey to ascertain if loss of smell and taste can be used as a bioindicator for asymptomatic COVID-19 carriers. It will also let us understand the cut-off for smell and taste sensitivity for carriers.

**Who can take this test?**

Anyone above 18 and having no prior allergy to some food items can take this test.

**How will you use my data?**

This is a research study and we will take the utmost care to protect the privacy of the participant and data.

**Is there any cost to participation?**

No, this is free of cost.

**What are the main items used for the test?**

These are some common household items e.g. lemon, mint, cumin, ghee, dhaniya, red-chilli, salt, sugar, vicks etc. There are multiple choices for the same categories of smell and taste. You can skip the individual tests or entire section, quit at any point of the test.

This app does not intend to replace any medical diagnosis. Please consult your doctor if you are experiencing any symptoms outlined here at WHO website.

### Would you like to test your smell and taste?

#### Do the Smell & Taste-Check!

You must be **18 years old or older** in order to participate.
You can take the Smell-&-Taste-Check as often as you like. We would like to encourage you to participate **approximately weekly** during the next 3 months.
Participation is voluntary and data is collected in pseudonymized fashion.

The ability to smell, taste and the sensitivity to irritation in the mouth and nose can be impaired for various reasons - e.g. in respiratory diseases by cold viruses and also by the new corona virus (SARS-CoV2; CoViD-19) or even by completely different diseases.

We would like to investigate how these senses fluctuate over time. The results will help to assess whether the senses are more or less affected in COVID-19 than in other diseases and healthy people.

Please read the information sheet on the study carefully. Among other things, it contains important information on data protection.

☐ I have read and understood the information sheet, I am over 18 years old, and I voluntarily participate.

#### Tell us a bit about yourself

Let's start with a few questions about you and your health.

Required questions are indicated with an asterisk *.

#### *1. In which year were you born?

*Mark only one oval.*

| | | | | |
|---|---|---|---|---|
| ◯ 2003 | ◯ 1982 | ◯ 1961 | ◯ 1940 | ◯ 1919 |
| ◯ 2002 | ◯ 1981 | ◯ 1960 | ◯ 1939 | ◯ 1918 |
| ◯ 2001 | ◯ 1980 | ◯ 1959 | ◯ 1938 | ◯ 1917 |
| ◯ 2000 | ◯ 1979 | ◯ 1958 | ◯ 1937 | ◯ 1916 |
| ◯ 1999 | ◯ 1978 | ◯ 1957 | ◯ 1936 | ◯ 1915 |
| ◯ 1998 | ◯ 1977 | ◯ 1956 | ◯ 1935 | ◯ 1914 |
| ◯ 1997 | ◯ 1976 | ◯ 1955 | ◯ 1934 | ◯ 1913 |
| ◯ 1996 | ◯ 1975 | ◯ 1954 | ◯ 1933 | ◯ 1912 |
| ◯ 1995 | ◯ 1974 | ◯ 1953 | ◯ 1932 | ◯ 1911 |
| ◯ 1994 | ◯ 1973 | ◯ 1952 | ◯ 1931 | ◯ 1910 |
| ◯ 1993 | ◯ 1972 | ◯ 1951 | ◯ 1930 | ◯ 1909 |
| ◯ 1992 | ◯ 1971 | ◯ 1950 | ◯ 1929 | ◯ 1908 |
| ◯ 1991 | ◯ 1970 | ◯ 1949 | ◯ 1928 | ◯ 1907 |
| ◯ 1990 | ◯ 1969 | ◯ 1948 | ◯ 1927 | ◯ 1906 |
| ◯ 1989 | ◯ 1968 | ◯ 1947 | ◯ 1926 | ◯ 1905 |
| ◯ 1988 | ◯ 1967 | ◯ 1946 | ◯ 1925 | ◯ 1904 |
| ◯ 1987 | ◯ 1966 | ◯ 1945 | ◯ 1924 | ◯ 1903 |
| ◯ 1986 | ◯ 1965 | ◯ 1944 | ◯ 1923 | ◯ 1902 |
| ◯ 1985 | ◯ 1964 | ◯ 1943 | ◯ 1922 | ◯ 1901 |
| ◯ 1984 | ◯ 1963 | ◯ 1942 | ◯ 1921 | ◯ 1900 |
| ◯ 1983 | ◯ 1962 | ◯ 1941 | ◯ 1920 | |

#### *2. Which gender do you most identify with?

*Mark only one oval.*

◯ Male
◯ Female
◯ Another gender not listed here
◯ Prefer not to say

*The following question pops up if 2nd or 3rd option is selected*

#### *3. Are you currently pregnant?

*Mark only one oval.*

- ◯ Yes
- ◯ No

#### *4. What is your current country of residence?

*Mark only one oval.*

- ◯ India
- ◯ Afghanistan
- ◯ Albania
- ◯ Algeria
- ◯ Andorra
- ◯ Angola
- ◯ Antigua and Barbuda
- ◯ Argentina
- ◯ Armenia
- ◯ Australia
- ◯ Austria
- ◯ Azerbaijan
- ◯ Bahamas
- ◯ Bahrain
- ◯ Bangladesh
- ◯ Barbados
- ◯ Belarus
- ◯ Belgium
- ◯ Belize
- ◯ Benin
- ◯ Bhutan
- ◯ Bolivia
- ◯ Bosnia and Herzegovina
- ◯ Botswana
- ◯ Brazil
- ◯ Brunei
- ◯ Bulgaria
- ◯ Burkina Faso
- ◯ Germany
- ◯ Ghana
- ◯ Greece
- ◯ Grenada
- ◯ Guatemala
- ◯ Guinea
- ◯ Guinea-Bissau
- ◯ Guyana
- ◯ Haiti
- ◯ Holy See
- ◯ Honduras
- ◯ Hungary
- ◯ Iceland
- ◯ Indonesia
- ◯ Iran
- ◯ Iraq
- ◯ Ireland
- ◯ Israel
- ◯ Italy
- ◯ Jamaica
- ◯ Japan
- ◯ Jordan
- ◯ Kazakhstan
- ◯ Kenya
- ◯ Kiribati
- ◯ Kuwait
- ◯ Kyrgyzstan
- ◯ Laos
- ◯ Oman
- ◯ Pakistan
- ◯ Palau
- ◯ Palestine State
- ◯ Panama
- ◯ Papua New Guinea
- ◯ Paraguay
- ◯ Peru
- ◯ Philippines
- ◯ Poland
- ◯ Portugal
- ◯ Qatar
- ◯ Romania
- ◯ Russia
- ◯ Rwanda
- ◯ Saint Kitts and Nevis
- ◯ Saint Lucia
- ◯ Saint Vincent and the Grenadines
- ◯ Samoa
- ◯ San Marino
- ◯ Sao Tome and Principe
- ◯ Saudi Arabia
- ◯ Senegal
- ◯ Serbia
- ◯ Seychelles
- ◯ Sierra Leone
- ◯ Singapore
- ◯ Slovakia

◯ Burundi
◯ Côte d'Ivoire
◯ Cabo Verde
◯ Cambodia
◯ Cameroon
◯ Canada
◯ Central African Republic
◯ Chad
◯ Chile
◯ China
◯ Colombia
◯ Comoros
◯ Congo (Congo-Brazzaville)
◯ Costa Rica
◯ Croatia
◯ Cuba
◯ Cyprus
◯ Czechia (Czech Republic)
◯ Democratic Republic of the Congo
◯ Denmark
◯ Djibouti
◯ Dominica
◯ Dominican Republic
◯ Ecuador
◯ Egypt
◯ El Salvador
◯ Equatorial Guinea
◯ Eritrea
◯ Estonia
◯ Eswatini (fmr. "Swaziland")
◯ Ethiopia
◯ Fiji
◯ Finland
◯ France
◯ Gabon
◯ Gambia
◯ Georgia
◯ Latvia
◯ Lebanon
◯ Lesotho
◯ Liberia
◯ Libya
◯ Liechtenstein
◯ Lithuania
◯ Luxembourg
◯ Madagascar
◯ Malawi
◯ Malaysia
◯ Maldives
◯ Mali
◯ Malta
◯ Marshall Islands
◯ Mauritania
◯ Mauritius
◯ Mexico
◯ Micronesia
◯ Moldova
◯ Monaco
◯ Mongolia
◯ Montenegro
◯ Morocco
◯ Mozambique
◯ Myanmar (formerly Burma)
◯ Namibia
◯ Nauru
◯ Nepal
◯ Netherlands
◯ New Zealand
◯ Nicaragua
◯ Niger
◯ Nigeria
◯ North Korea
◯ North Macedonia
◯ Norway
◯ Slovenia
◯ Solomon Islands
◯ Somalia
◯ South Africa
◯ South Korea
◯ South Sudan
◯ Spain
◯ Sri Lanka
◯ Sudan
◯ Suriname
◯ Sweden
◯ Switzerland
◯ Syria
◯ Tajikistan
◯ Tanzania
◯ Thailand
◯ Timor-Leste
◯ Togo
◯ Tonga
◯ Trinidad and Tobago
◯ Tunisia
◯ Turkey
◯ Turkmenistan
◯ Tuvalu
◯ Uganda
◯ Ukraine
◯ United Arab Emirates
◯ United Kingdom
◯ United States of America
◯ Uruguay
◯ Uzbekistan
◯ Vanuatu
◯ Venezuela
◯ Vietnam
◯ Yemen
◯ Zambia
◯ Zimbabwe

**5. What state, city, or town do you live in?**

Enter state, city, town here_____________

**6. What is your zip / postal code?**

Enter zip code here___________________

***7. What is your highest level of formal education? E.g. school, vocational training, university, etc.**

*Mark only one oval.*

- ◯ No formal education
- ◯ 10th
- ◯ 12th
- ◯ Bachelors
- ◯ Masters or above

***8. How many contacts do you have every day outside of your household? E.g. through work, public transport, etc.**

*Mark only one oval.*

- ◯ none
- ◯ <5
- ◯ 5-10
- ◯ 11-20
- ◯ 21-50
- ◯ >50

***9. Have you been diagnosed with or suspected you had a respiratory illness in the past 14 days? (e.g COVID-19, flu, cold, etc)**

*Mark only one oval.*

- ◯ Yes
- ◯ No

***10. Have you been diagnosed with COVID-19, in the past 14 days?**

*Mark only one oval.*

- ◯ No-do not have any symptoms
- ◯ Yes-diagnosed by medical symptoms only
- ◯ Yes-diagnosed with viral swab

◯ Yes-diagnosed with another lab test
◯ Unknown-I was tested but did not get my results
◯ No-not diagnosed, but have symptoms
◯ No-had a negative test, but still have symptoms

#### 11. When date did you first notice symptoms of your COVID illness? Provide your best guess or leave blank if you do not remember.

Click here to select date

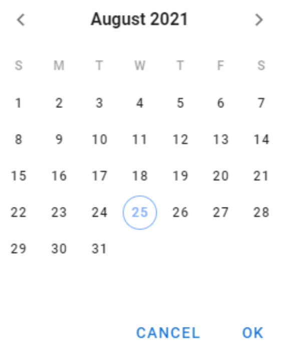


#### *12. Have you been diagnosed with or suspected with any other respiratory illness (not COVID-19)?

◯ Bacterial illness (e.g, strep throat)
◯ Viral illness (e.g, flu/common cold)
◯ Other respiratory illness
◯ I don't know
◯ None of the above

#### 13. When date did you first notice symptoms of your other respiratory illness? Provide your best guess or leave blank if you do not remember.

Click here to select date

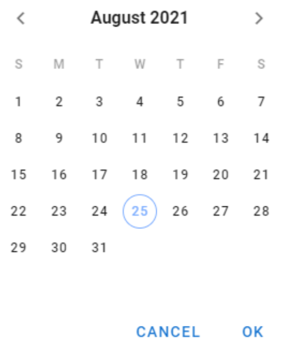

***14. <u>In the past 14 days</u>, have you had any of the following symptoms?**

◯ Fever
◯ Dry Cough
◯ Cough with mucus
◯ Difficulty breathing/shortness of breath
◯ Chest tightness
◯ Running nose
◯ Sore throat
◯ Changes in food flavour
◯ Changes in smell
◯ Changes in taste
◯ Loss of appetite
◯ Headache
◯ Body aches
◯ Fatigue
◯ Diarrhea
◯ Abdominal pain
◯ Nausea
◯ Skin sesitivity
◯ Dry mouth
◯ No symptoms

***15. Have you experienced any changes of smell in the past 14 days?**

*Mark only one oval.*

◯ Slight
◯ Moderate
◯ Complete Loss
◯ None

***16. Have you experienced any changes to any of the specific taste in the past 14 days?**

◯ Sweet
◯ Sour
◯ Salty
◯ Bitter
◯ None

***17. Have you ever been a regular smoker, e-cigarette user, or vaper, in your life?**

*Mark only one oval.*

◯ Yes-former smoker
◯ Yes-current smoker
◯ No
◯ Prefer not to say

**18. <u>In the last 6 months</u>, have you had any of the following?**

◯ High blood pressure
◯ Heart diseases (heart attack)
◯ Diabetes (high blood sugar)
◯ Obesity
◯ Lung diseases (asthma/COPD)
◯ Head trauma
◯ Neurological diseases
◯ Cancer that required chemotherapy or radiation
◯ Chronic sinus problems
◯ Smell disorder (e.g, hyposmia/anosmia)
◯ Taste disorder (e.g, dysgeusia)
◯ Seasonal allergies/hay fever
◯ Dry mouth
◯ Other
◯ None

SMELL TASTE TRACKING

**Thank you!**

### Get ready for the Smell & Taste Check!

Important notice!! Please pay attention

In 4 short parts, you can measure your sense of smell and taste and other sensations in your nose and mouth. It should be done at home because you will smell and taste 7 different items in your household. It will take approximately 10-15 minutes.

**Preparations:** Try to avoid distracting smells like scented lotions or perfumes. Perform the check in a room with minimal smells - like those from air fresheners, cigarette smoke, or cooking. Do not smoke, eat or drink anything (other than drinking water) for at least 30 min prior.

**Attention:**
This assessment does not replace medical testing, and it does not provide a medical diagnosis. Please do not take any specific action, medical or otherwise, based on the results of this self-assessment.

**Do not use any items you might have an allergy or sensitivity to. If you suffer from a medical condition such as asthma that makes it hard for you to sniff several odors in succession, you may skip the odor section. Please skip if you are unsure whether your medical condition allows you to participate.**
You may skip items. But we would like to encourage you be consistent throughout repeated checks.

SMELL
TASTE
TRACKING

**Part 1: Smell**

You will need 7 of the odor items listed on the next page. If possible, please try them at room temperature, and in a state where they are most intense.

For example, an unpeeled banana, apple will not smell very strongly. Peeling, slicing, or crushing items may help release their smell

*** Required selection of ingredient**
From each of the 7 groups below, please select and gather one item that is normally found in your household.

Select those items that will be available over the coming weeks. If you repeat the check, **please try not to switch items.**

If you suffer from a medical condition such as asthma that makes it hard for you to sniff several odors in succession, **you may skip the odor section.**

☐

I would like to skip the smell/odor section.

Choose available items below. Once done, press NEXT to start with odor/smell test.

***scented cosmetics or detergents:**

Click button on left and right to browse through ingredients

Selected item: laundry detergent

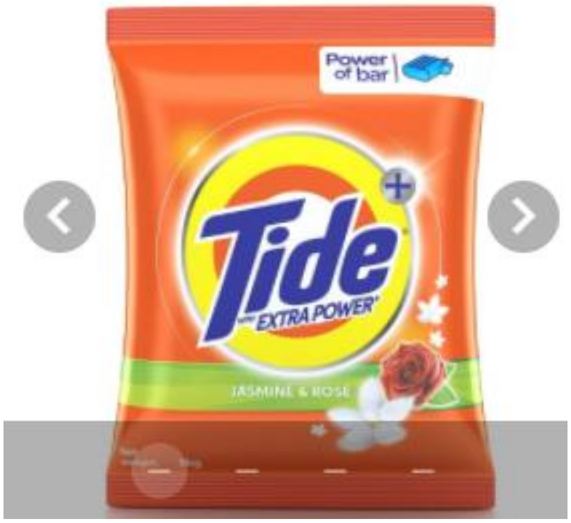


Selected item: shampoo or conditioner

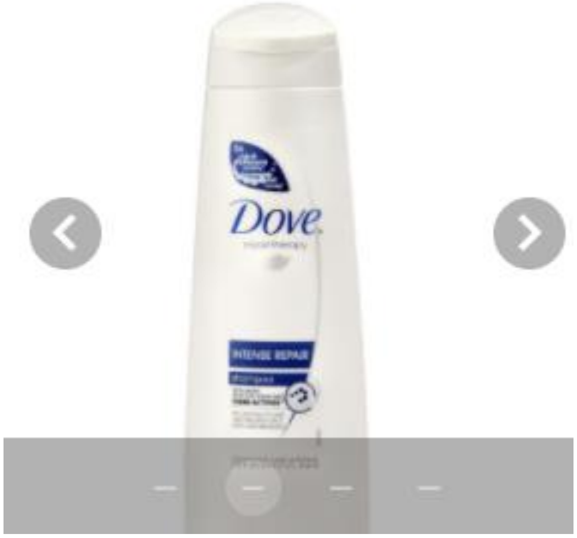


Selected item: scented soap

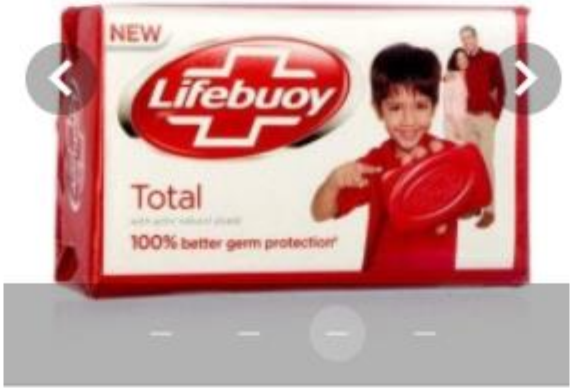


Selected item: None of above is available

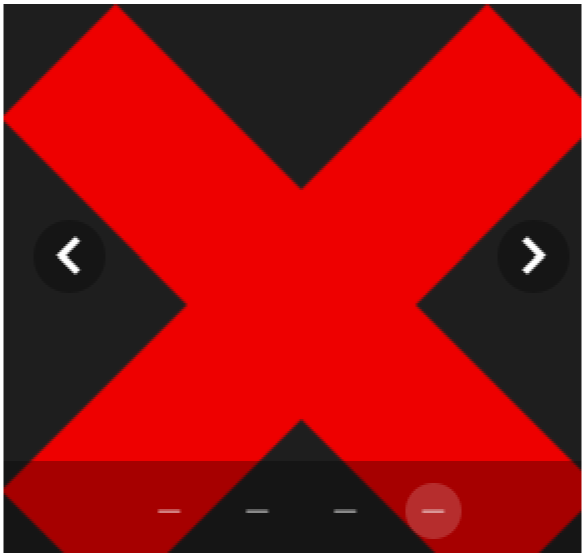

***spices, herbs or food ingredients1**

Click button on left and right to browse through ingredients

Selected item: Cinnamon(Dalchini)

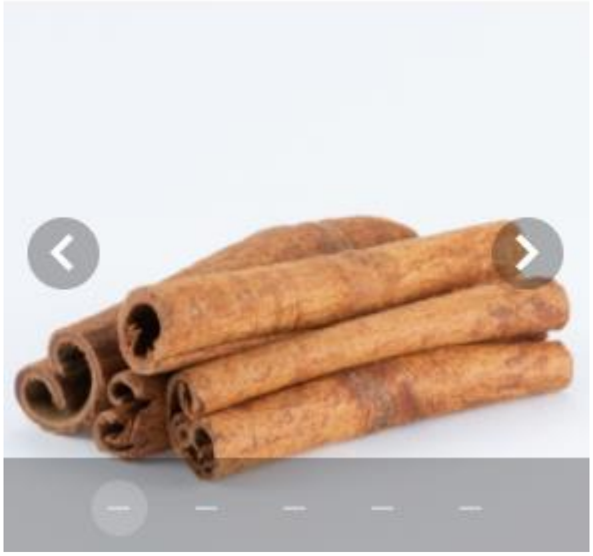

Selected item: Clove(lavang)

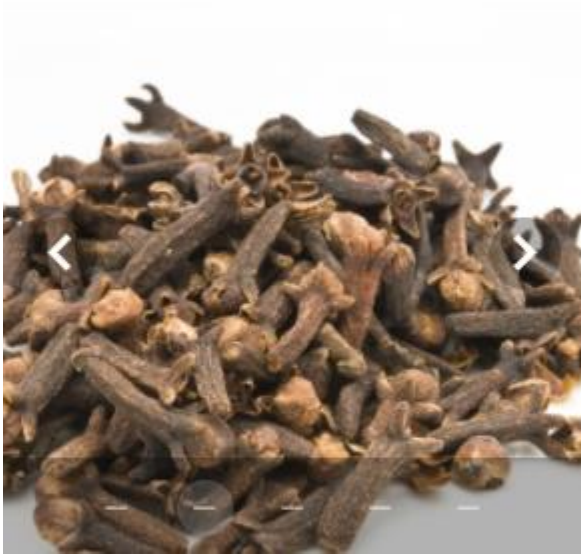

Selected item: Cardamom (Elaichi)

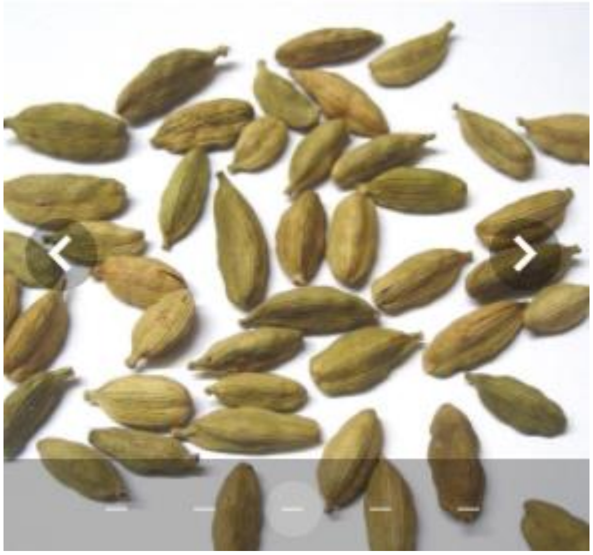

Selected item: Nutmeg(jaifal)

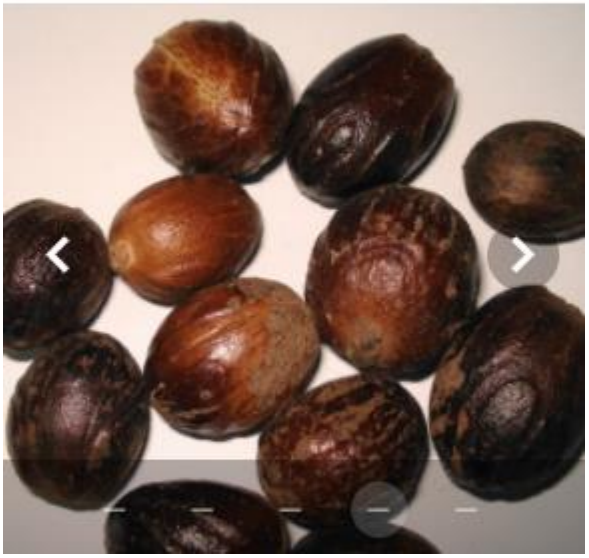

Selected item: None of above is available

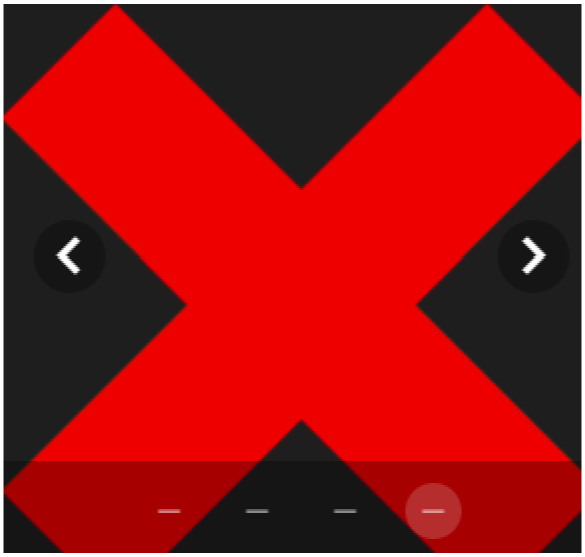

***spices, herbs or food ingredients2**

Click button on left and right to browse through ingredients

Selected item: Cumin/Jeera

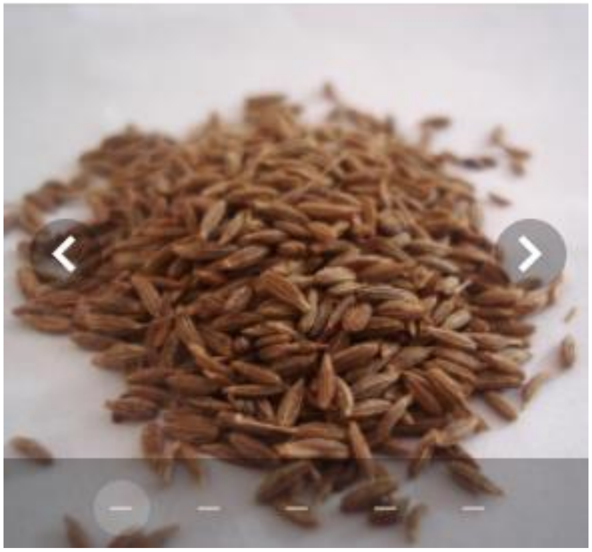

Selected item: Ajwain/Carom seeds

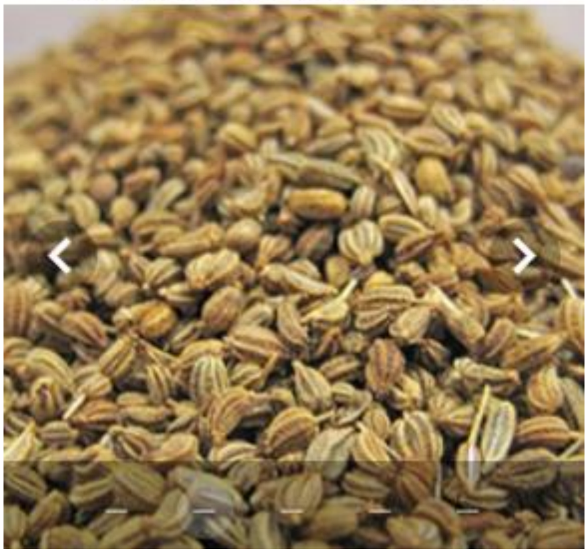

Selected item: Fennel/Saunf

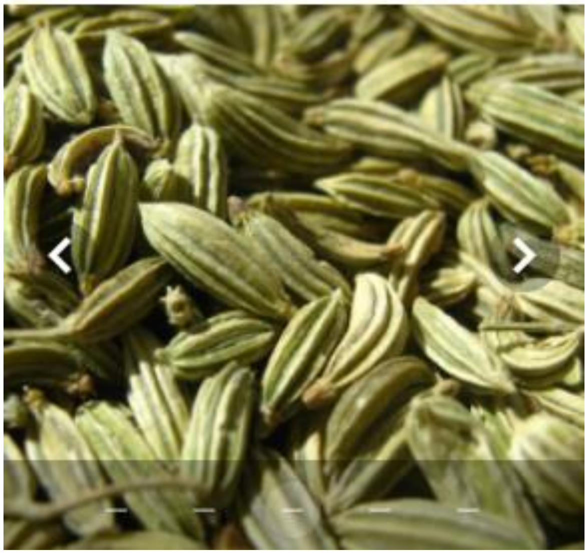

Selected item: Coriander seeds/Dhania

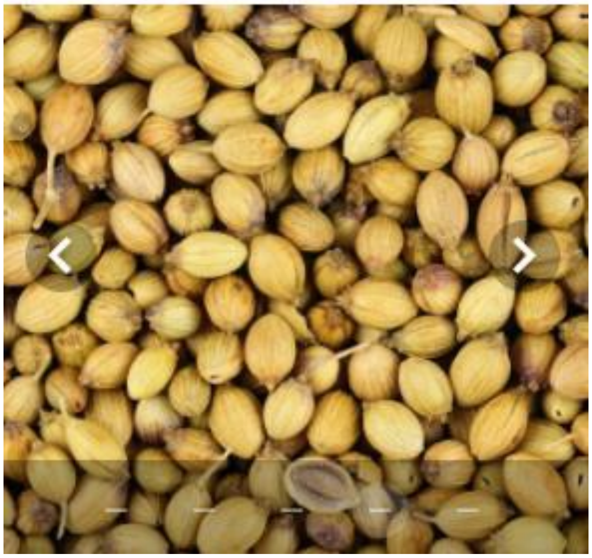

Selected item: None of above is available

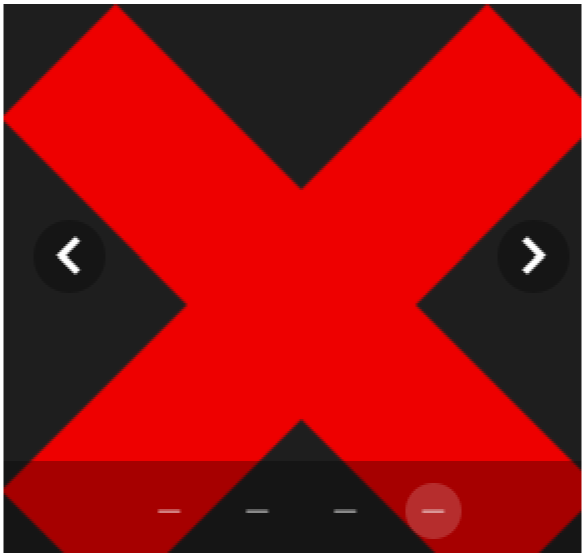

***spice mixtures**

Click button on left and right to browse through ingredients

Selected item: Garam/Goda masala

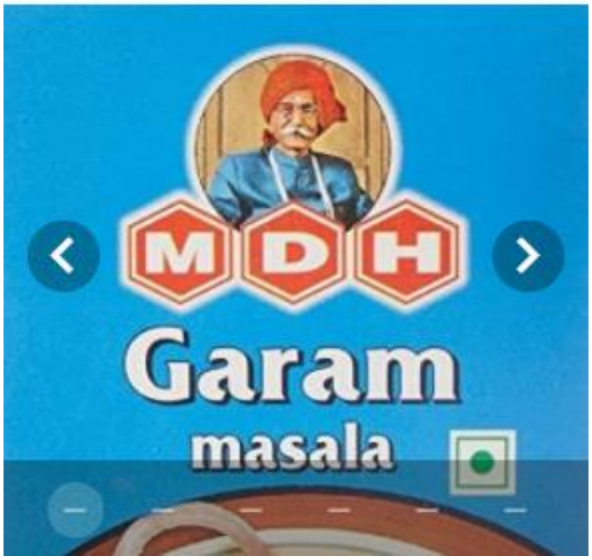


Selected item: Sambhar/Rasam/Puliyogare powder

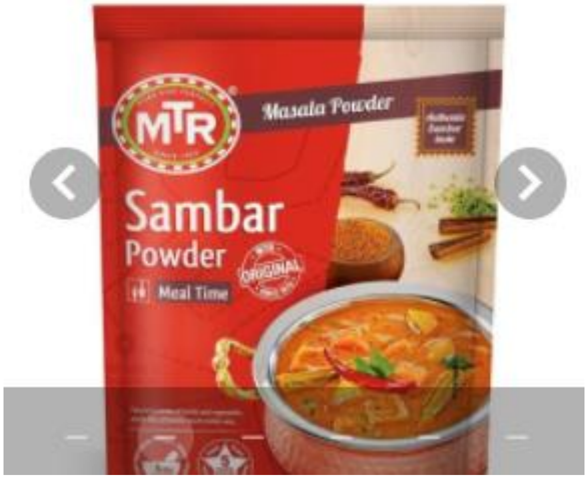


Selected item: Biryani/Fish/Chicken/Meat masala

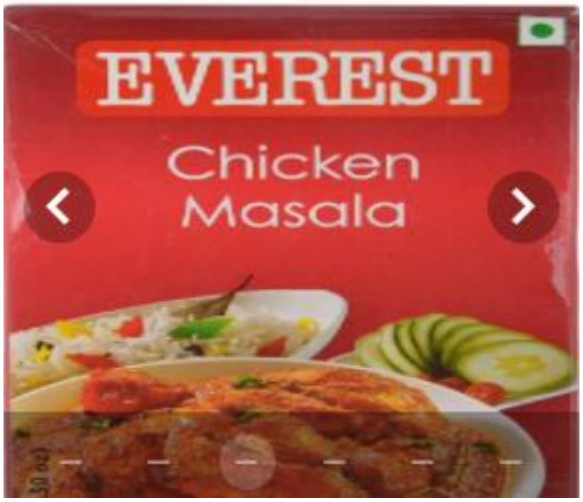


Selected item: Maggi masala

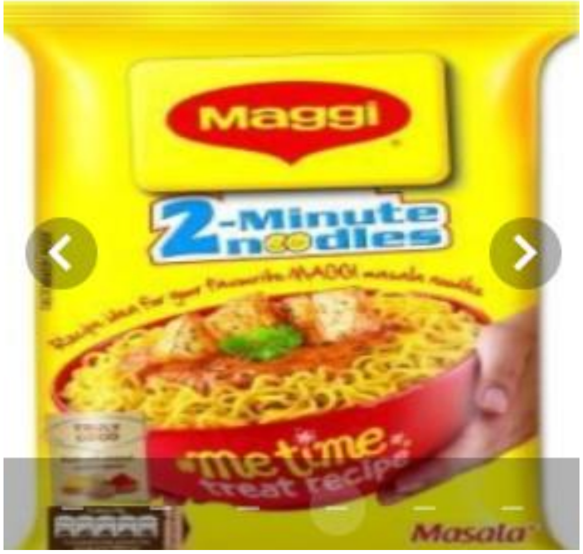

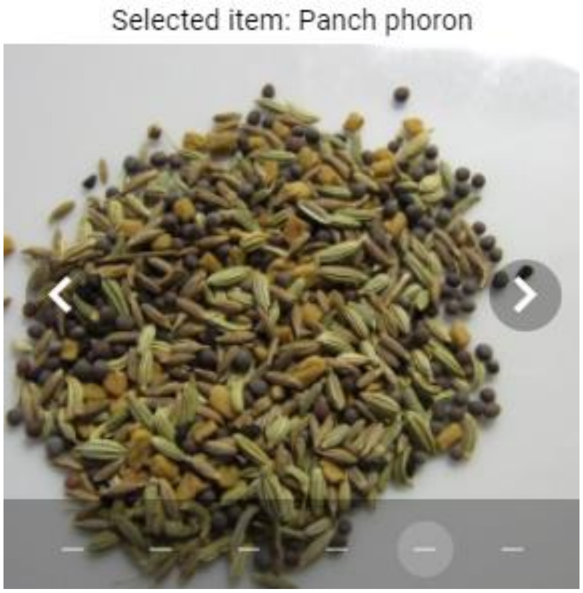


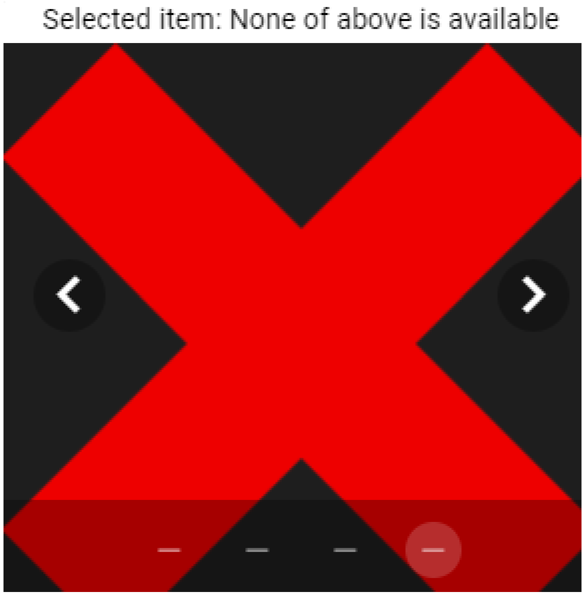


***fruits, fruit juice or vegetables**

Click button on left and right to browse through ingredients

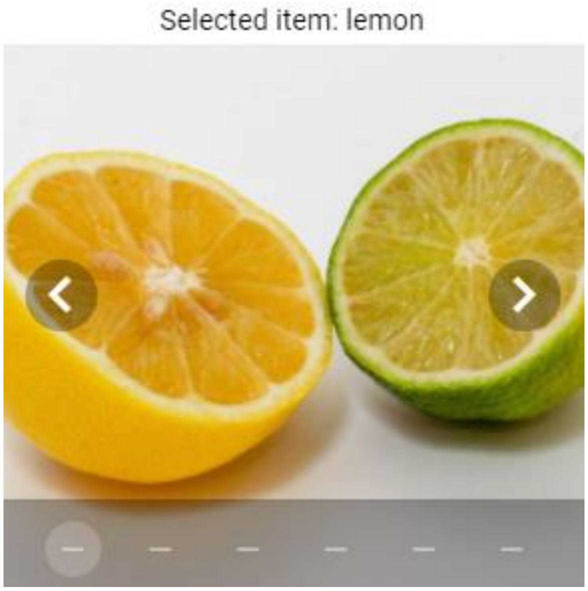


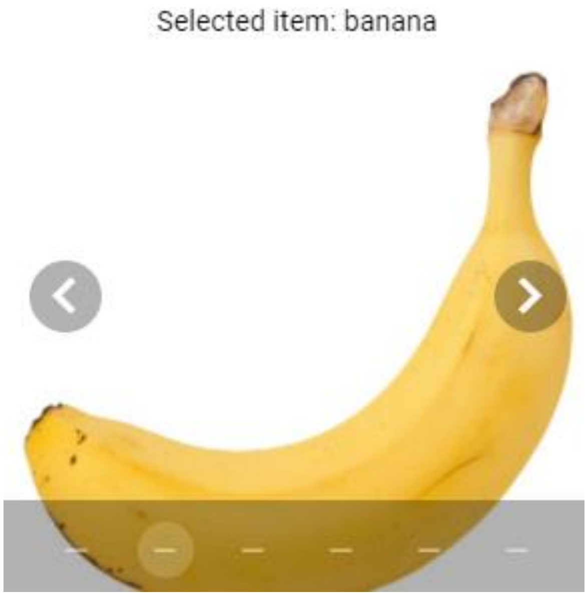

Selected item: apple

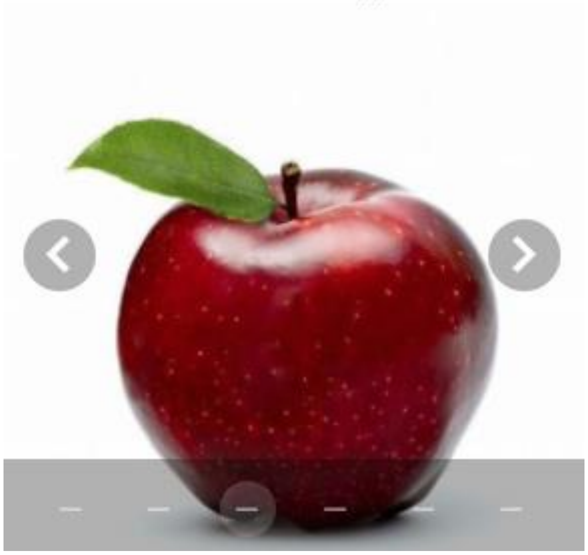

Selected item: cucumber

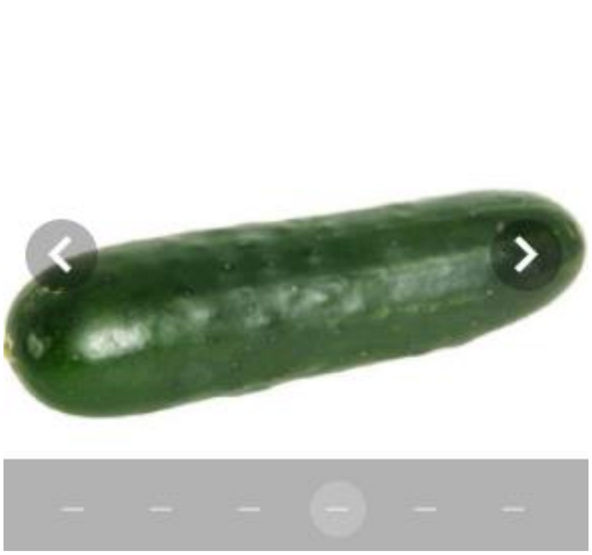

Selected item: tomato

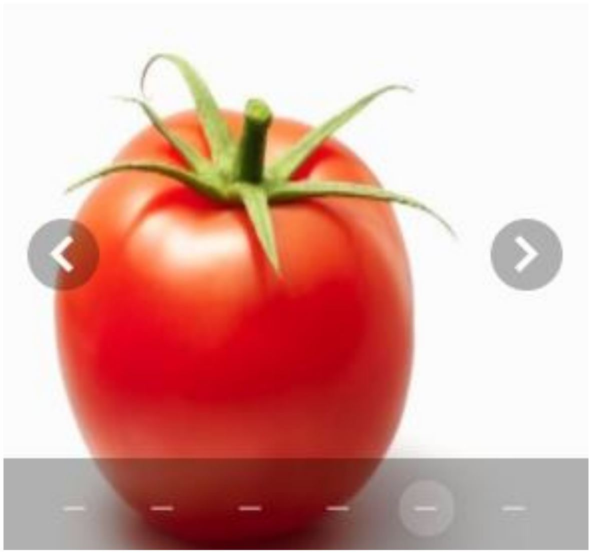

Selected item: None of above is available

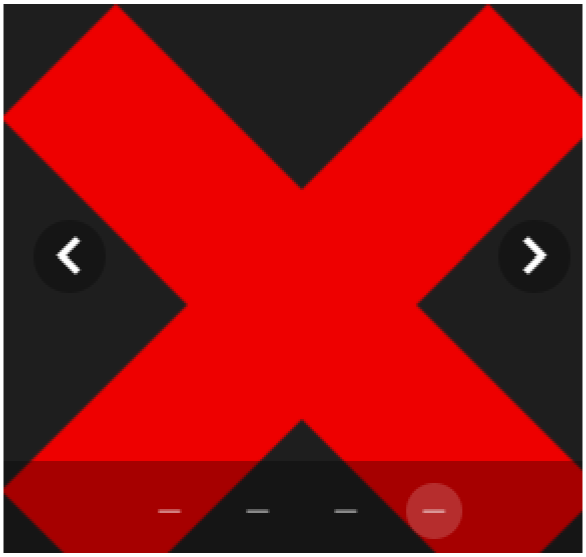

***Dairy items:**

Click button on left and right to browse through ingredients

Selected item: Milk

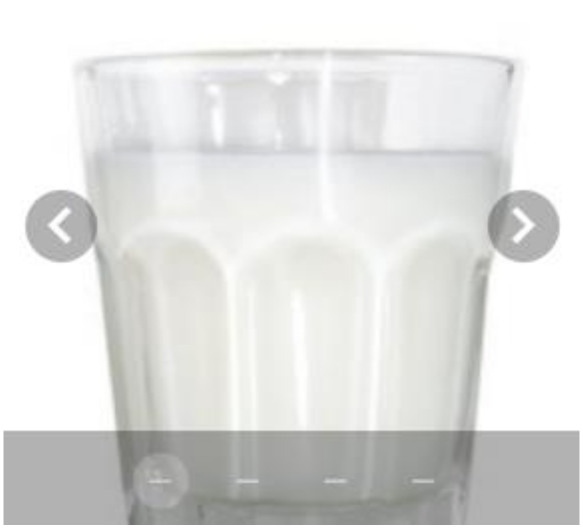

Selected item: Ghee

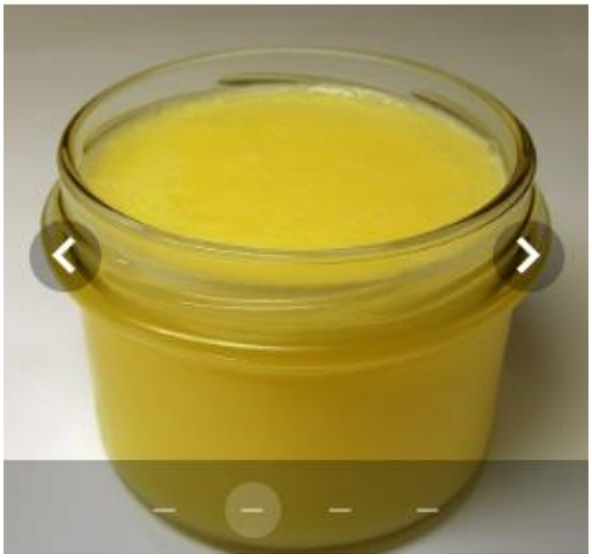

Selected item: Butter

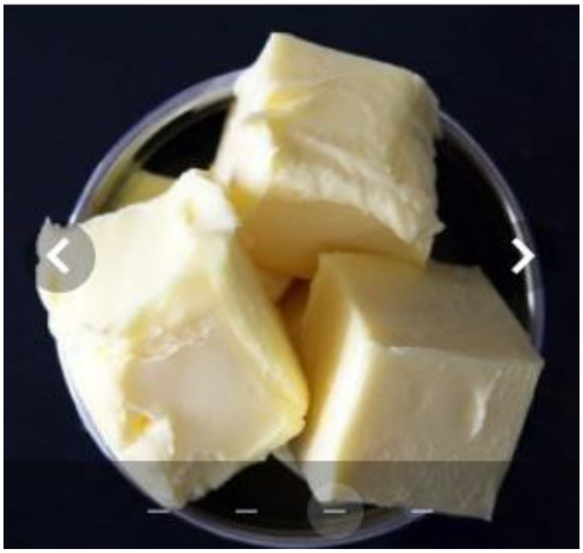

Selected item: None of above is available

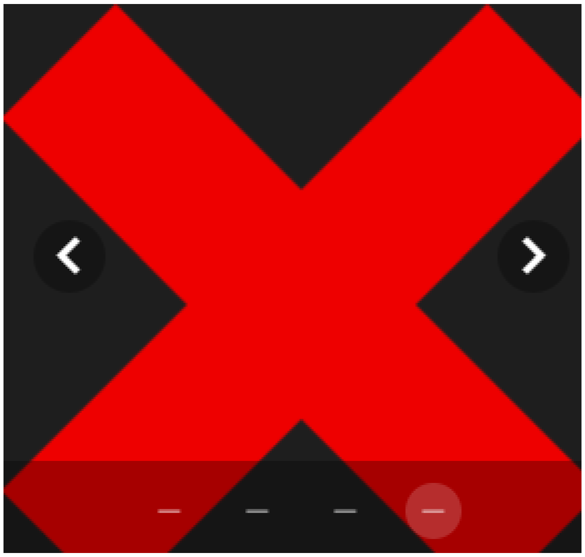

Selected item: coffee

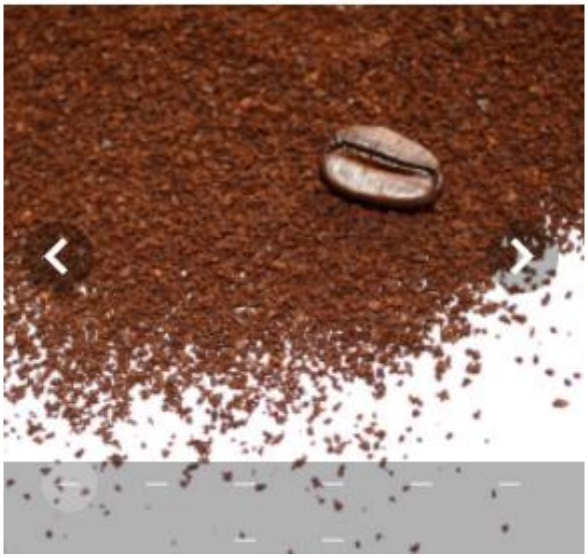

Selected item: burnt matches

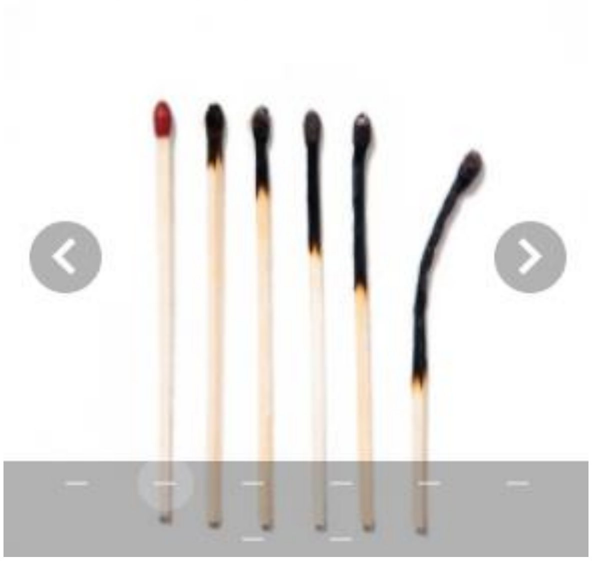

Selected item: cigarette butts or ashtray

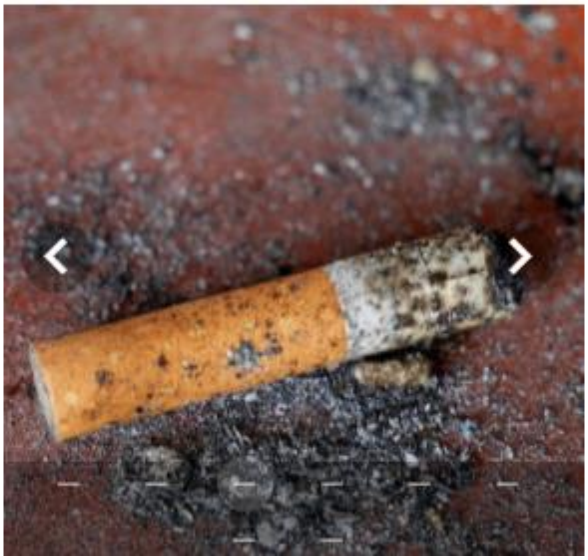

Selected item: chocolate

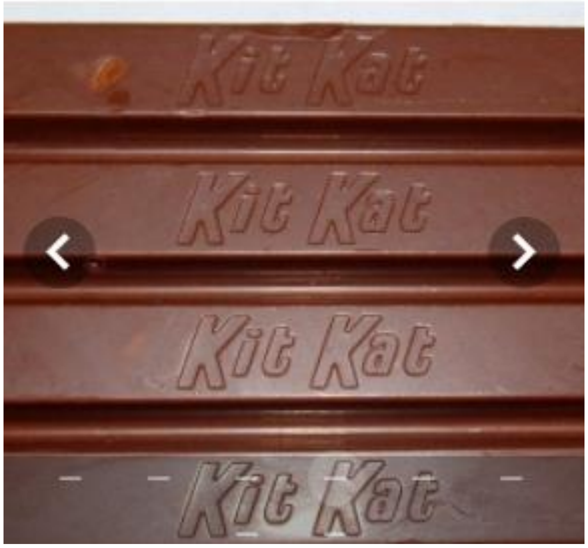

Selected item: crushed grass/leaves

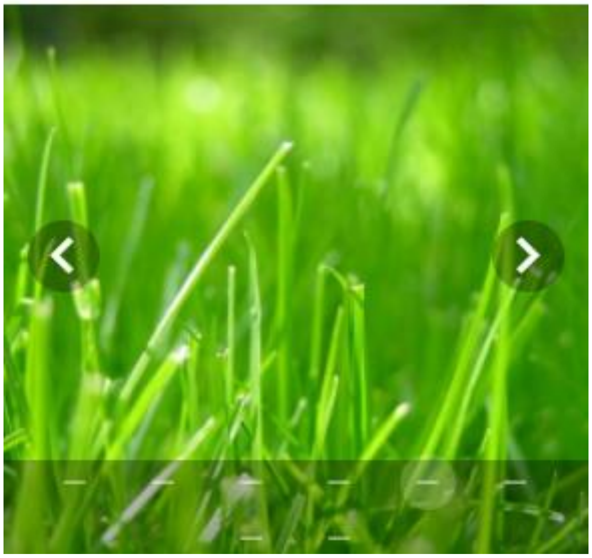

Selected item: wet soil

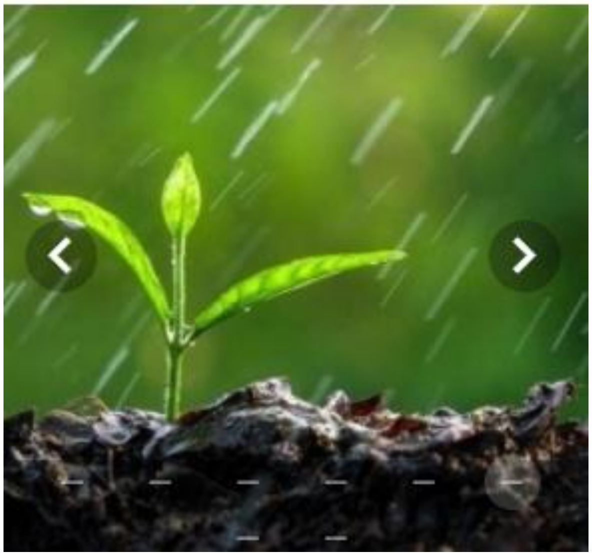

Selected item: tea leaves

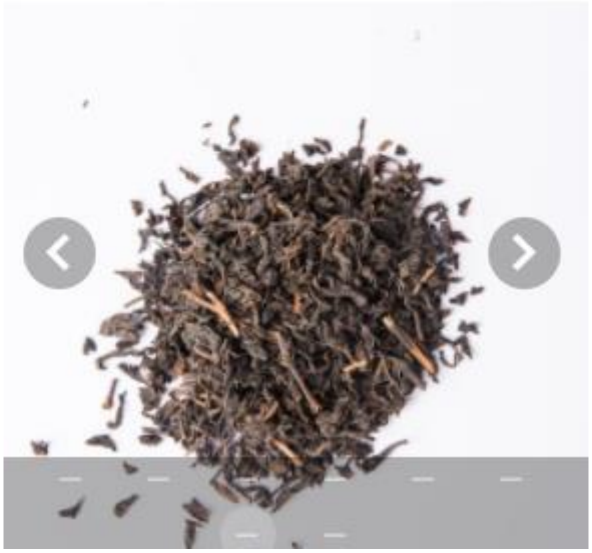

Selected item: None of above is available

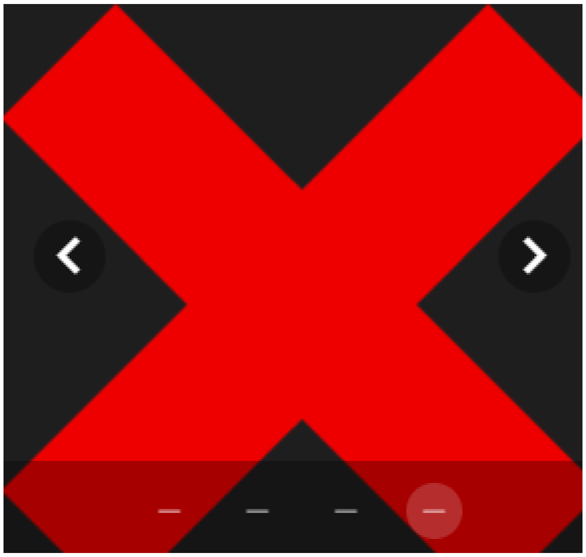

#### Part 1 of 4: Smell

*** Required question**

To smell, bring the objects selected earlier to your nose one by one and sniff normally 1-3 times. Here you can see how it's done.

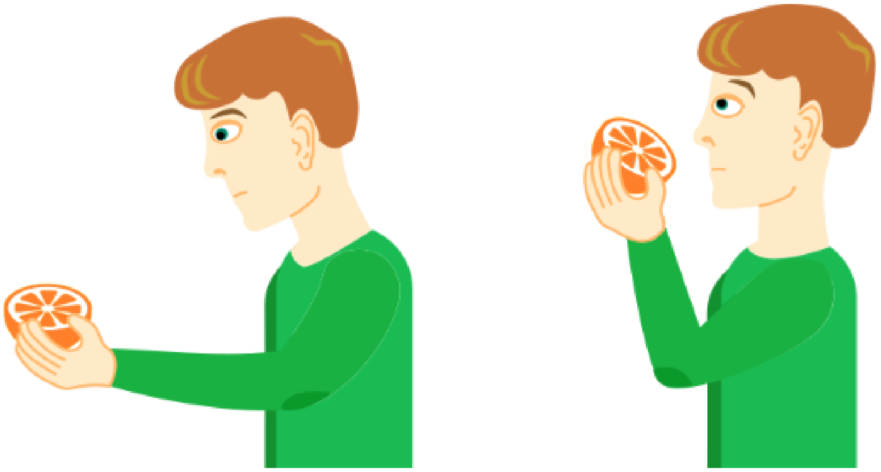

Once you are done sniffing, rate the smell intensity below

**(If you cannot smell the item at all, please move the slider all of the way to the left.)**

***19. Rate the smell for <Scented Detergents>**

No sensation — Very intense

***20. Rate the smell for <Spices/Herbs-1>**

No sensation — Very intense

***21. Rate the smell for <Spices/Herbs-2>**

No sensation — Very intense

***22. Rate the smell for <Spice Mixtures>**

No sensation — Very intense

***23. Rate the smell for <Fruits/Vegetables>**

No sensation — Very intense

***24. Rate the smell for <Dairy items>**

No sensation — Very intense

***25. Rate the smell for <Other items>**

No sensation — Very intense

SMELL
TASTE
TRACKING

#### Part 2:
#### Other sensations in your nose.

#### Part 2 of 4: Other sensations in your nose

*** Required question**
Please select and gather 1 item from the list below. You will be asked to smell it and rate the intensity of the cooling, tickling, stinging, or burning sensation.

***Please select and gather 1 of the following items:**
Click button on left and right to browse through ingredients

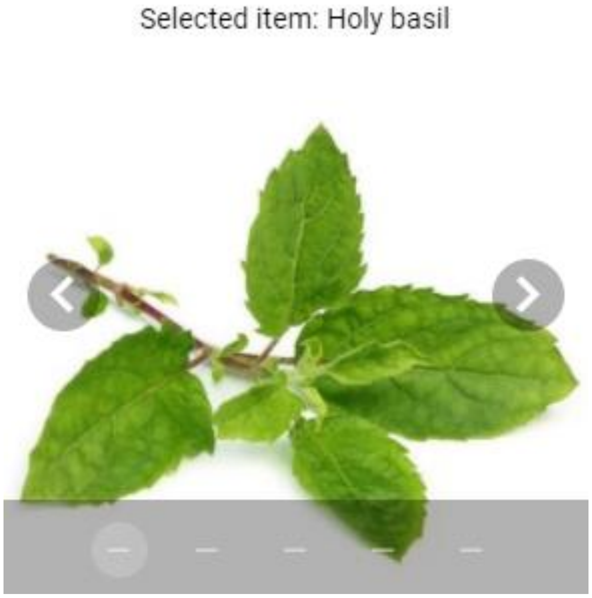


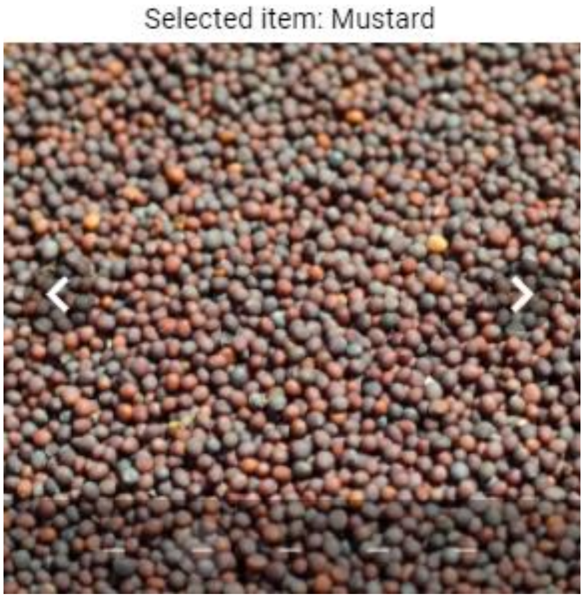

Selected item: Mint leaves

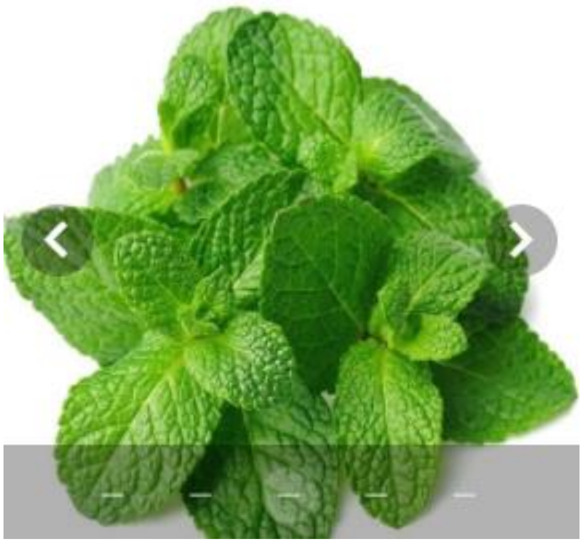

Selected item: Vicks Vaporub or Camphor

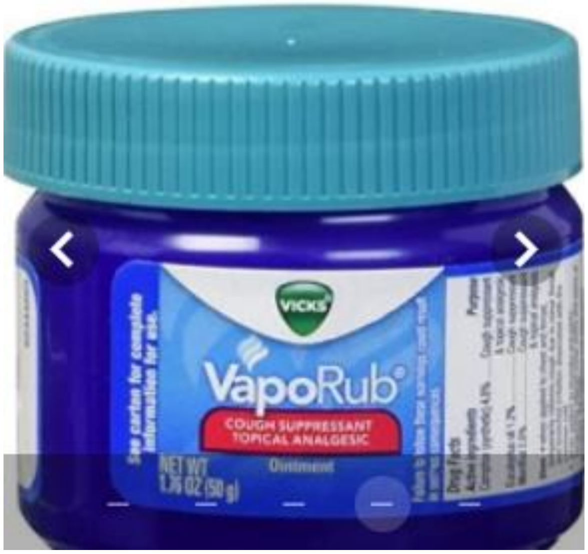


Selected item: None of above is available

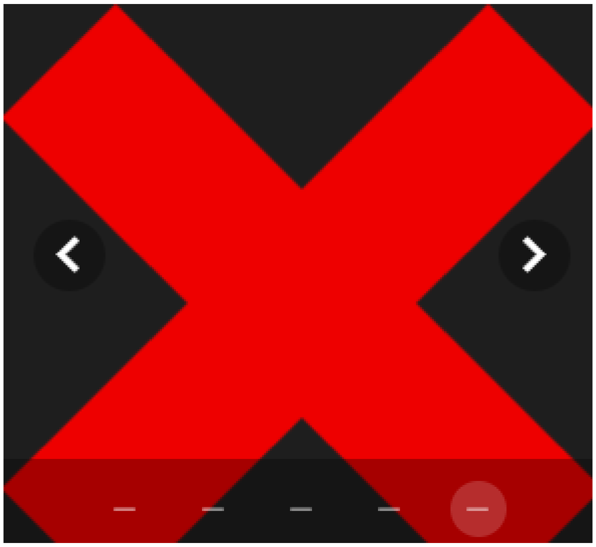

To smell, bring the item to your nose and sniff normally 1-3 times.

Once you are done sniffing, rate the **cooling, tickling, burning or stinging sensation** below
**(If you cannot smell the item at all, please move the slider all of the way to the left.)**

***26. Rate the smell for the object selected above.**

No sensation Very intense

SMELL
TASTE
TRACKING

**Part 3: Taste**
**(sweet, sour, salty, bitter)**

#### Part 3 of 4: Taste (sweet, sour, salty, bitter)

Please select and gather 4 items from the list below. You will be asked to taste them and rate the intensity of the taste.

Please gather the following four items:

***Sweet:**

Click button on left and right to browse through ingredients

Selected item: Sugar

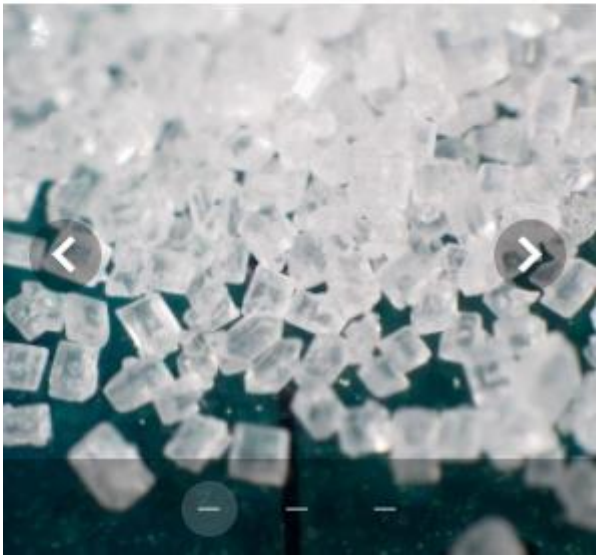

Selected item: Sweetener

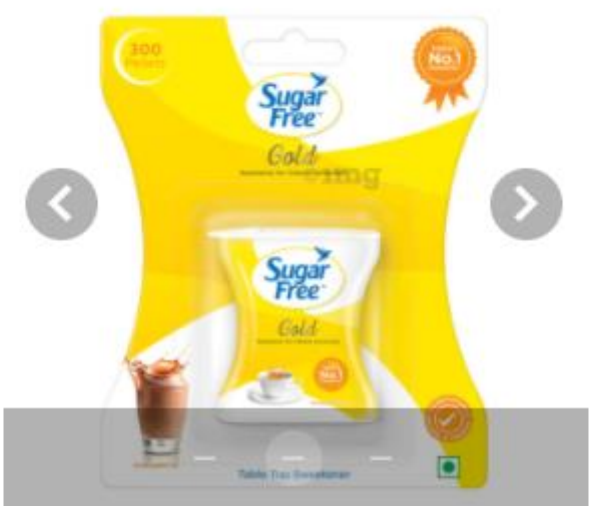

Selected item: None of above is available

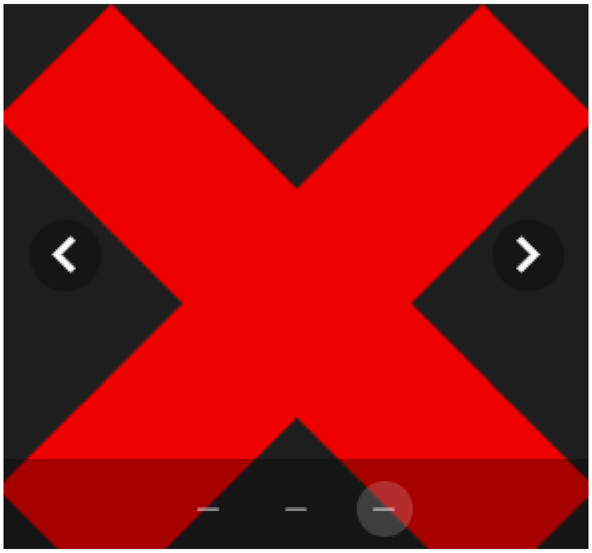

***Salty:**

Click button on left and right to browse through ingredients

Selected item: Salt

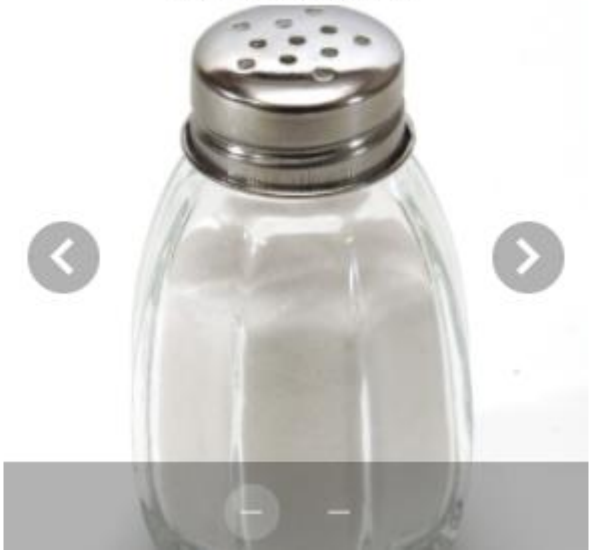

Selected item: None of above is available

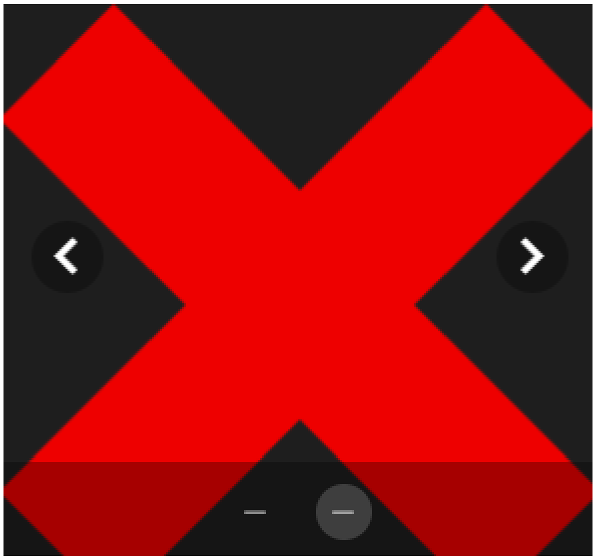

***Sour:**

Click button on left and right to browse through ingredients

Selected item: lemon juice

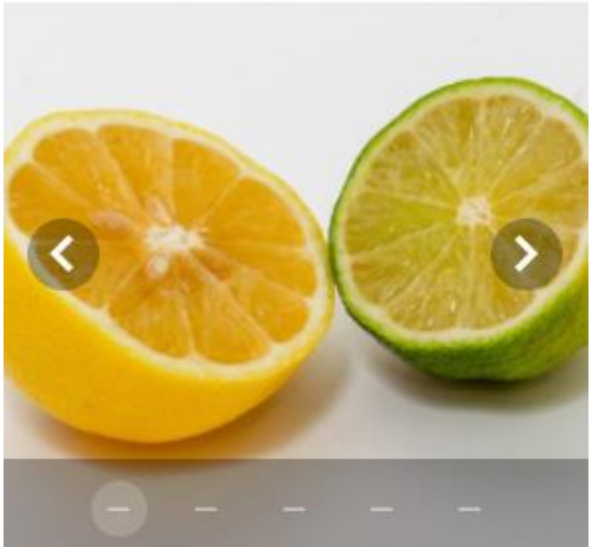

Selected item: Tamarind

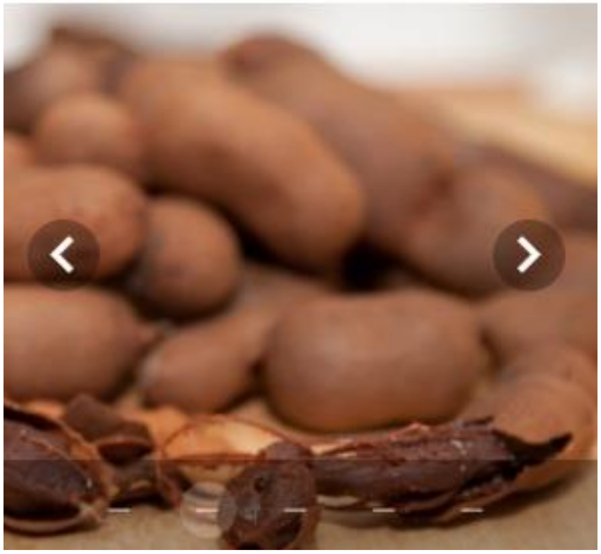

Selected item: Kokam

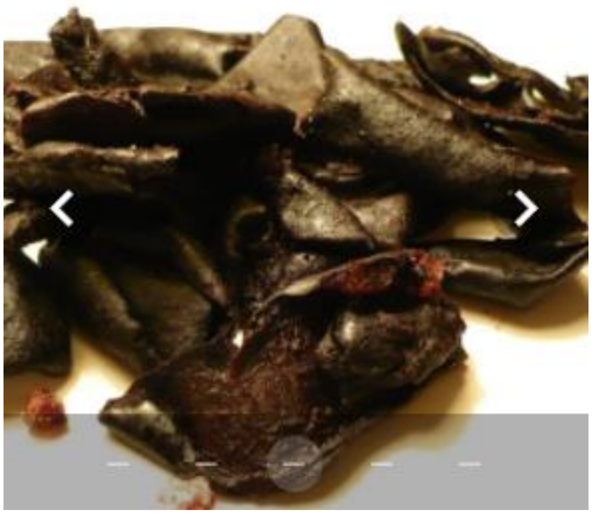

Selected item: Vinegar (any kind but balsamic)

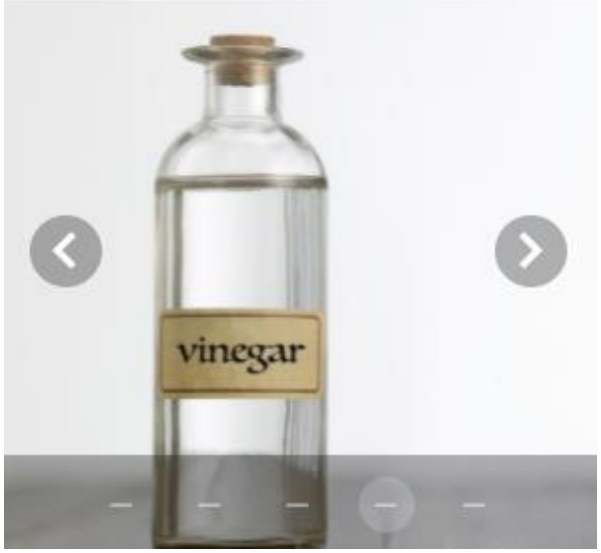

Selected item: None of above is available

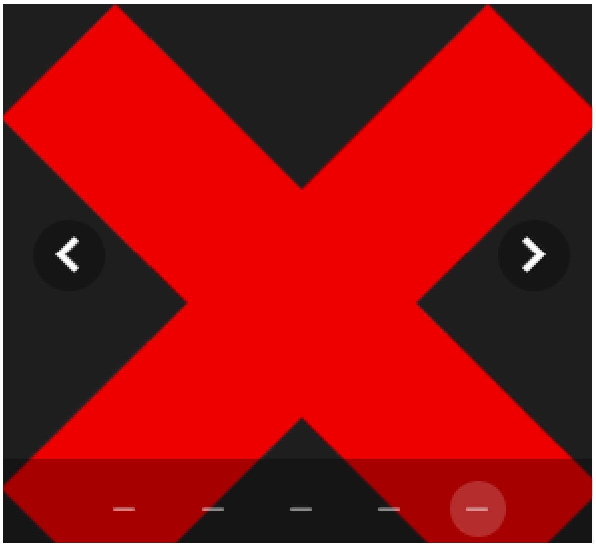

***Bitter:**

Click button on left and right to browse through ingredients

Selected item: black or green tea leaves/grains/dust from tea bags

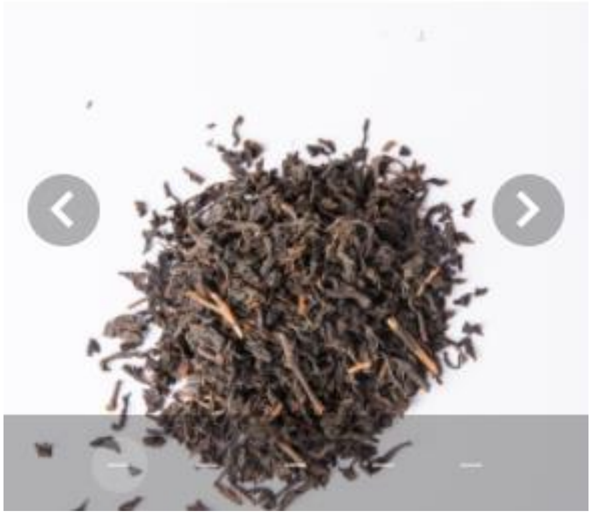

Selected item: Coffee (beans, grounds, instant powder)

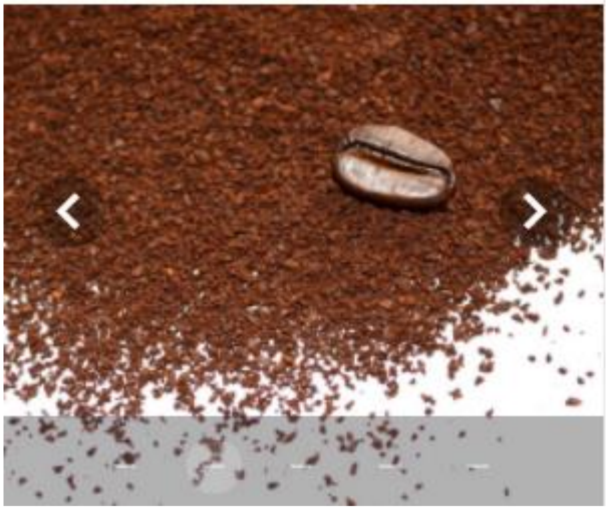

Selected item: Neem leaves

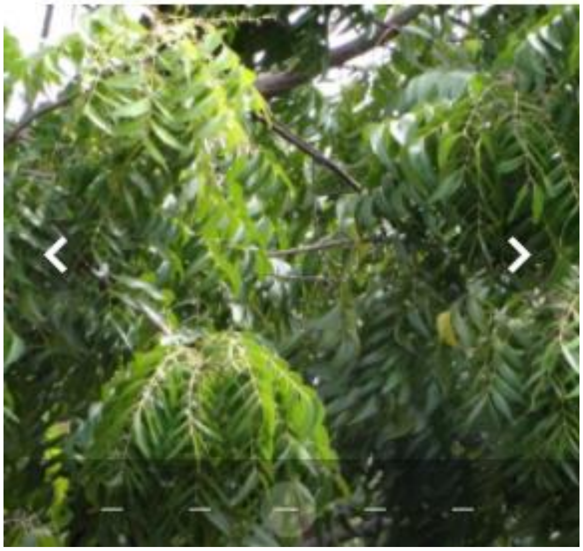

Selected item: Fenugreek

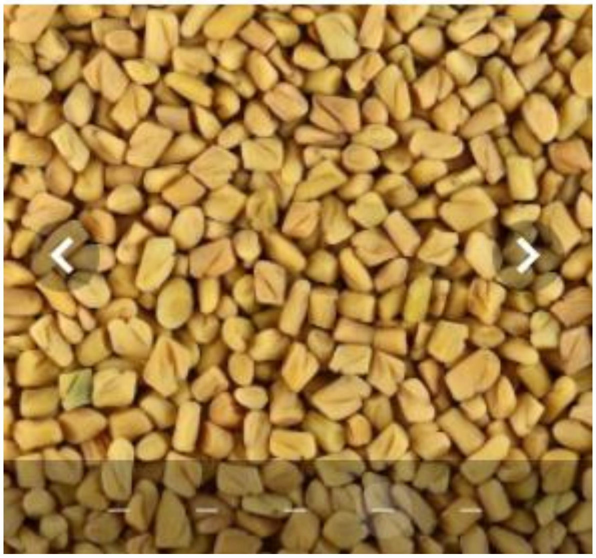

Selected item: None of above is available

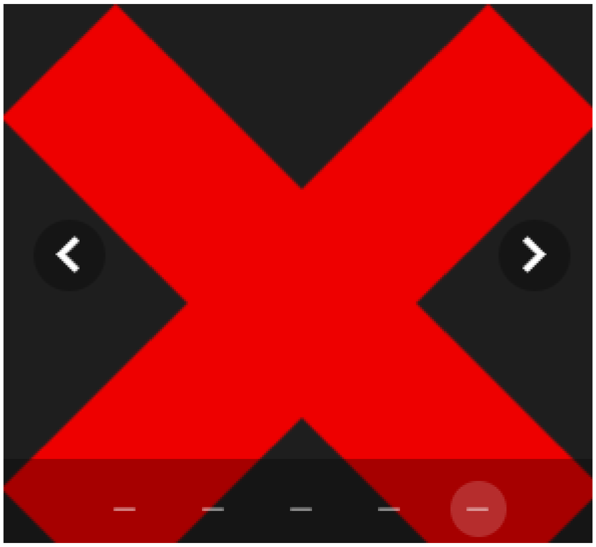

**Please also gather a teaspoon and a glass of water. Once gathered, please press NEXT**

#### Part 3 of 4: Taste (sweet, sour, salty, bitter)

**Take a small sip of water and rinse your mouth.**
Then, put a little less than a ¼ teaspoon of **black or green tea leaves/grains/dust from tea bags** on your tongue, close your mouth and move it around in your mouth.

***27. Rate the sweet taste intensity**

No sensation ... Very intense

***28. Rate the salty taste intensity**

No sensation ... Very intense

***29. Rate the sour taste intensity**

No sensation ... Very intense

***30. Rate the bitter taste intensity**

No sensation ... Very intense

SMELL
TASTE
TRACKING

**Part 4:**
**Other sensations in your mouth**

For example, burning from chili, cooling from mint, or tingling from carbonated beverages.

#### Part 4 of 4: Other sensations in your mouth

Please select and gather 1 item from the list below. You will be asked to place it in your mouth and rate the intensity of the cooling, tickling, stinging, or burning sensation.

Please select and gather 1 of the following items:
Click button on left and right to browse through ingredients

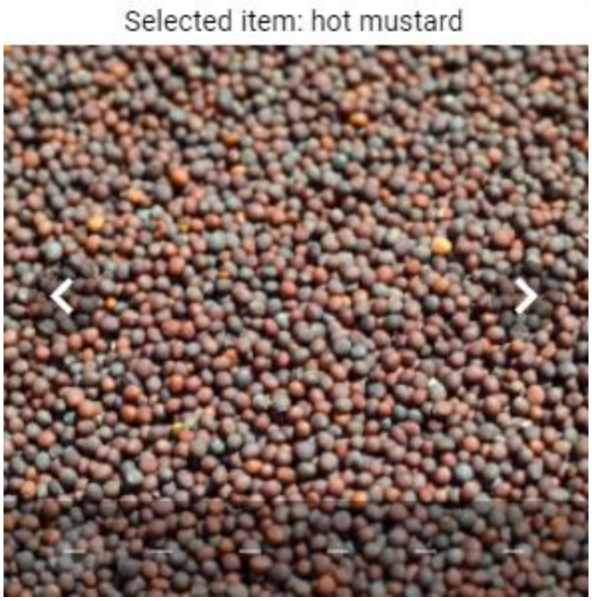


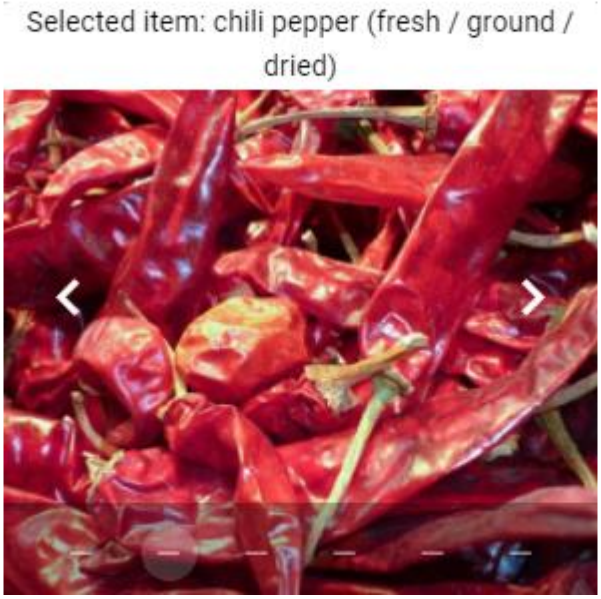

Selected item: Black Pepper

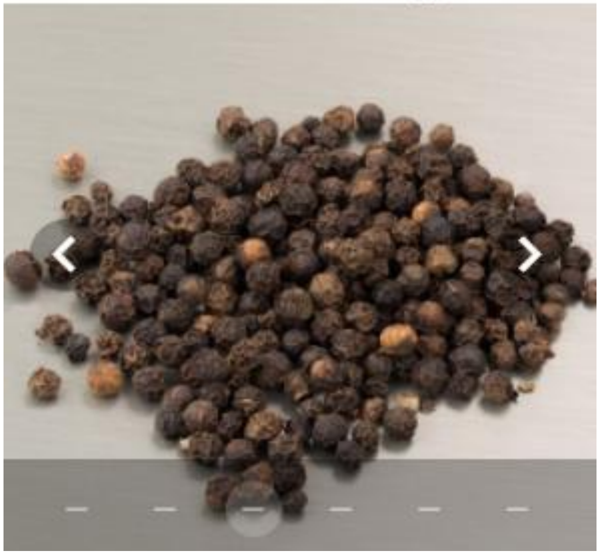

Selected item: ENO (normal sip)

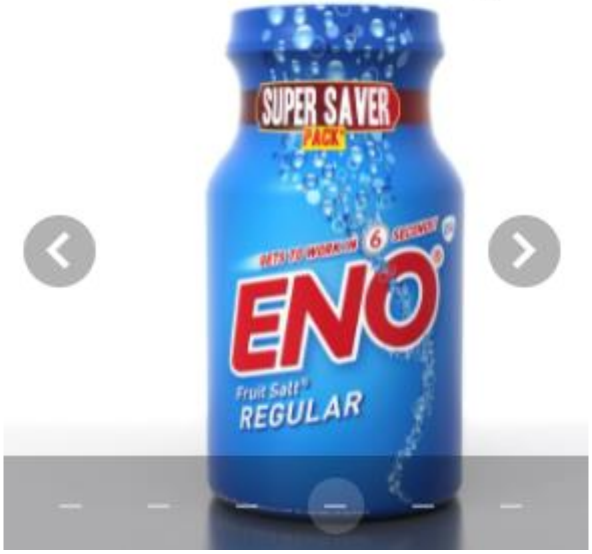


Selected item: mint leaves

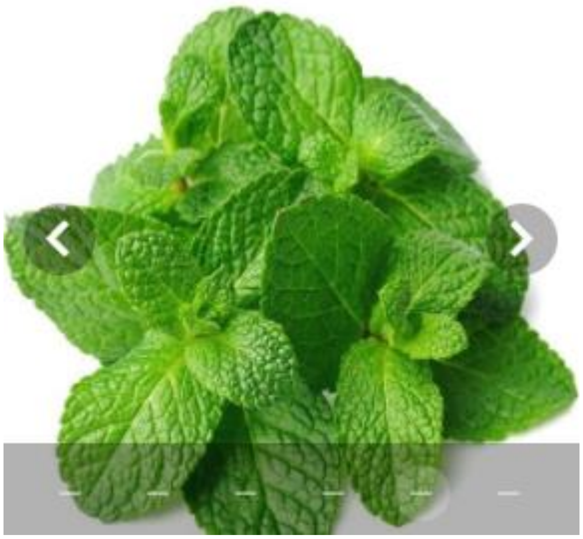

Selected item: None of above is available

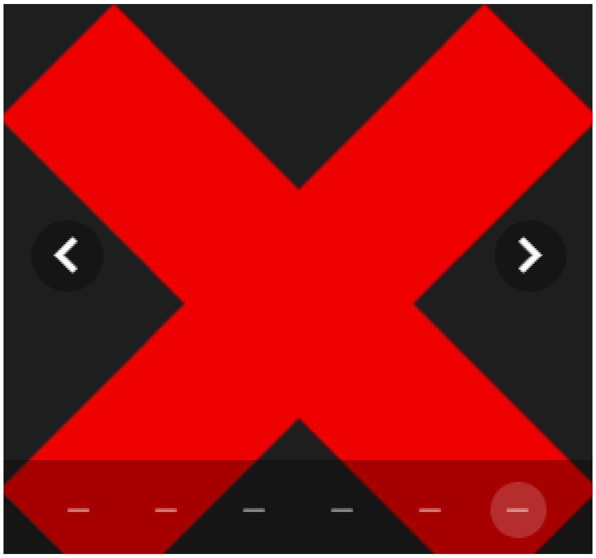

***31. Rate the cooling, tickling, burning or stinging sensation.**

No sensation — Very intense

Thank you for taking the smell and taste survey. We strongly recommend you to the report any change in smell and taste along with COVID status on this website.

**Please submit your response now.**

SUBMIT FORM

### Sample Response

Your responses indicate that you are experiencing notable taste change in the past 2 weeks. It is observed that there is a reduced sense of smell or taste in approximately 30-60% of COVID-19 cases even in the absence of other typical symptoms. It is strongly recommended that you consider self-quarantine along with continued monitoring of taste and smell loss on this website.

To further track your smell and taste change in the coming weeks, please REGISTER with us.

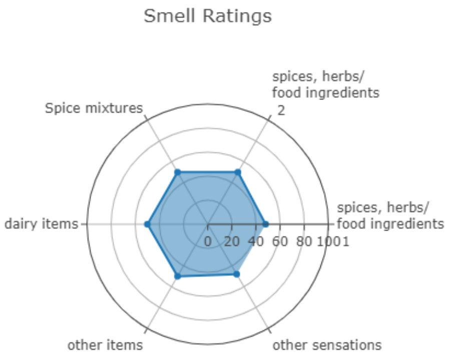


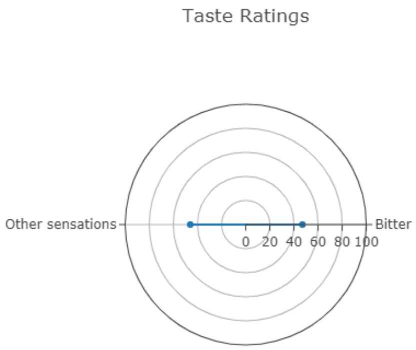


REGISTER FILL ANOTHER QUESTIONNAIRE

This app does not intend to replace any medical diagnosis. Please consult your doctor if you are experiencing any symptoms outlined at the WHO website

It is strongly encouraged to report any changes in sense of smell and taste along with any future COVID-19 testing and results on this website. It will be help us in determining that if smell and taste sensitivity can identify asymptomatic carriers. We will also be able to determine a cut-off for smell and taste sensitivity that indicates likely infection.
