## Supplementary Material A for "Leveraging machine learning and self-administered tests to predict COVID-19: An olfactory and gustatory dysfunction assessment through crowd-sourced data in India"

### SUPPLEMENTARY MATERIAL (PART A)

**Table S1.** The optimal parameters for the five machine learning models trained on five different sets of features. For readability, only parameters with non-default values are shown. (GEN-Generic, OBSAT-Olfactory Based Self Administered Tests and GBSAT-Gustatory Based Self Administered Tests.)

| Features | Model | Optimal Parameters |
|---|---|---|
| **GEN** | **Naive Bayes** | var_smoothing=0.0123 |
| | **Decision Tree** | max_depth=1, min_samples_split=5, random_state=0 and class_weight="balanced" |
| | **Random Forest** | criterion="entropy", max_depth=10, max_features="log2", min_samples_split=5, n_estimators=200, random_state=0 and class_weight="balanced" |
| | **Logistic Regression** | max_iter=500, solver="liblinear", random_state=0 and class_weight="balanced" |
| | **Support Vector Machines** | gamma=0.5, probability=True, random_state=0 and class_weight="balanced" |
| **OBSAT** | **Naive Bayes** | var_smoothing=0.4329 |
| | **Decision Tree** | max_depth=10, min_samples_split=5, random_state=0 and class_weight="balanced" |
| | **Random Forest** | max_depth=10, min_samples_split=10, random_state=0 and class_weight="balanced" |
| | **Logistic Regression** | max_iter=500, fit_intercept=False penalty="l1", solver="liblinear", random_state=0 and class_weight="balanced" |
| | **Support Vector Machines** | gamma=0.0001, probability=True, random_state=0 and class_weight="balanced" |
| **GBSAT** | **Naive Bayes** | var_smoothing=0.23101 |
| | **Decision Tree** | max_depth=10, min_samples_split=10, random_state=0 and class_weight="balanced" |
| | **Random Forest** | max_depth=5, random_state=0 and class_weight="balanced" |
| | **Logistic Regression** | C=100, max_iter=500, penalty="l1", solver="liblinear", fit_intercept=False, random_state=0 and class_weight="balanced" |
| | **Support Vector Machines** | C=10, gamma=0.01, probability=True, random_state=0 and class_weight="balanced" |
| **OBSAT + GBSAT** | **Naive Bayes** | var_smoothing=0.4329 |
| | **Decision Tree** | max_depth=5, min_samples_split=100, random_state=0 and class_weight="balanced" |
| | **Random Forest** | max_depth=5, random_state=0 and class_weight="balanced" |
| | **Logistic Regression** | C=10, max_iter=500, penalty="l1", solver="liblinear", random_state=0 and class_weight="balanced" |
| | **Support Vector Machines** | gamma=0.0001, probability=True, random_state=0 and class_weight="balanced" |
| **GEN + OBSAT + GBSAT** | **Naive Bayes** | var_smoothing=0.0187 |
| | **Decision Tree** | max_depth=1, min_samples_split=5, random_state=0 and class_weight="balanced" |
| | **Random Forest** | max_depth=10, min_samples_split=10, criterion="entropy" random_state=0 and class_weight="balanced" |
| | **Logistic Regression** | max_iter=500, penalty="l1", solver="liblinear", random_state=0 and class_weight="balanced" |
| | **Support Vector Machines** | gamma=0.1, probability=True, random_state=0 and class_weight="balanced" |

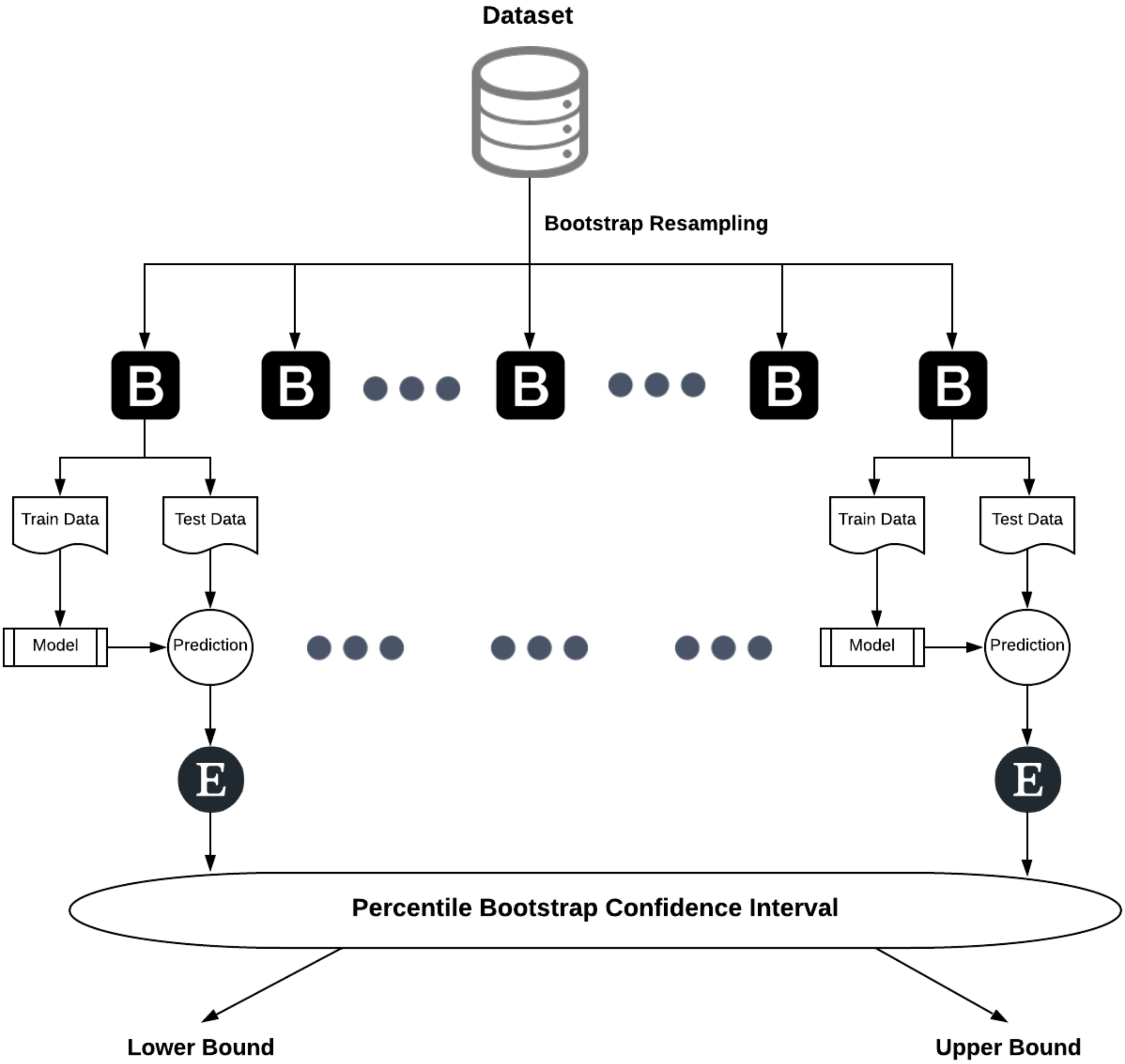


**Figure S1.** Assessing machine learning models through bootstrapping with resampling. Overall one thousand bootstraps (B) were generated using resampling method. All the bootstraps were then divided into train and test data for assessing the prediction capabilities using various evaluation metrics (E). Using the scores from all the bootstraps, lower and upper bound values were finally generated with 95% confidence interval.

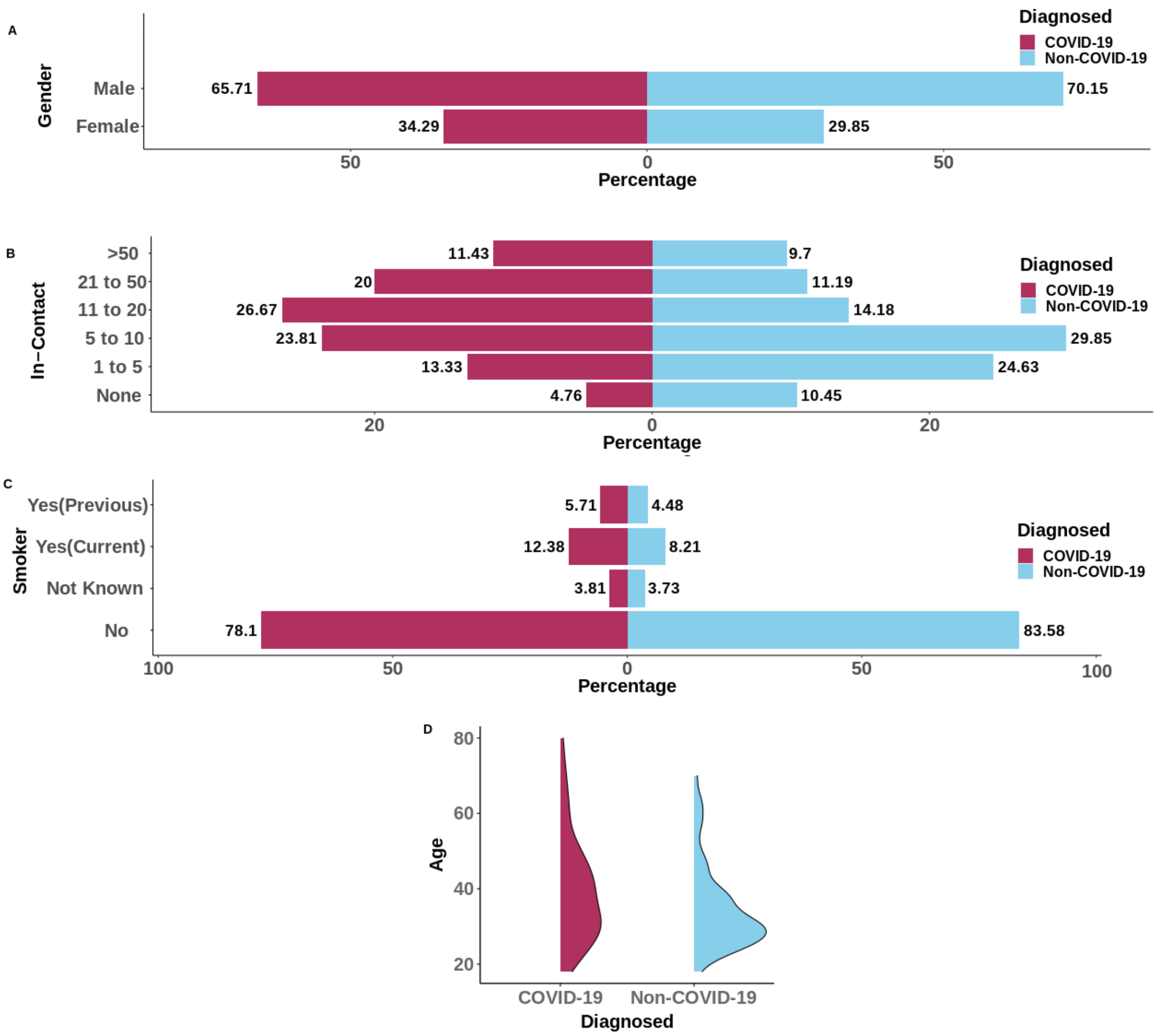


**Figure S2.** Comparison of demographic features i.e gender (A) and age (D), along with two other features i.e. Smoker (C) and In-Contact(B) between participants diagnosed as COVID-19 and Non-COVID-19.

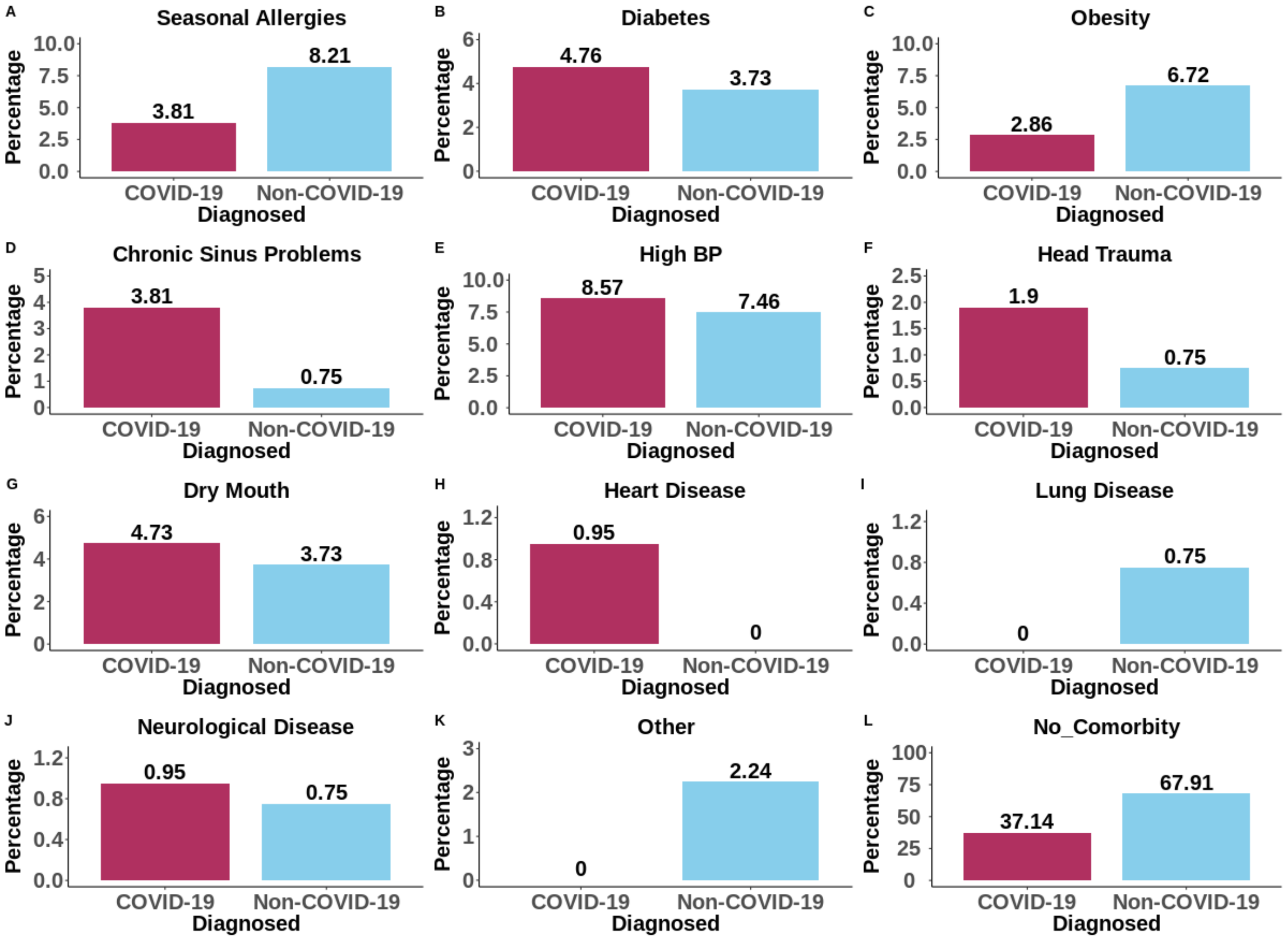


**Figure S3.** Reported health issues (faced in the last six months) of participants with and without COVID-19.

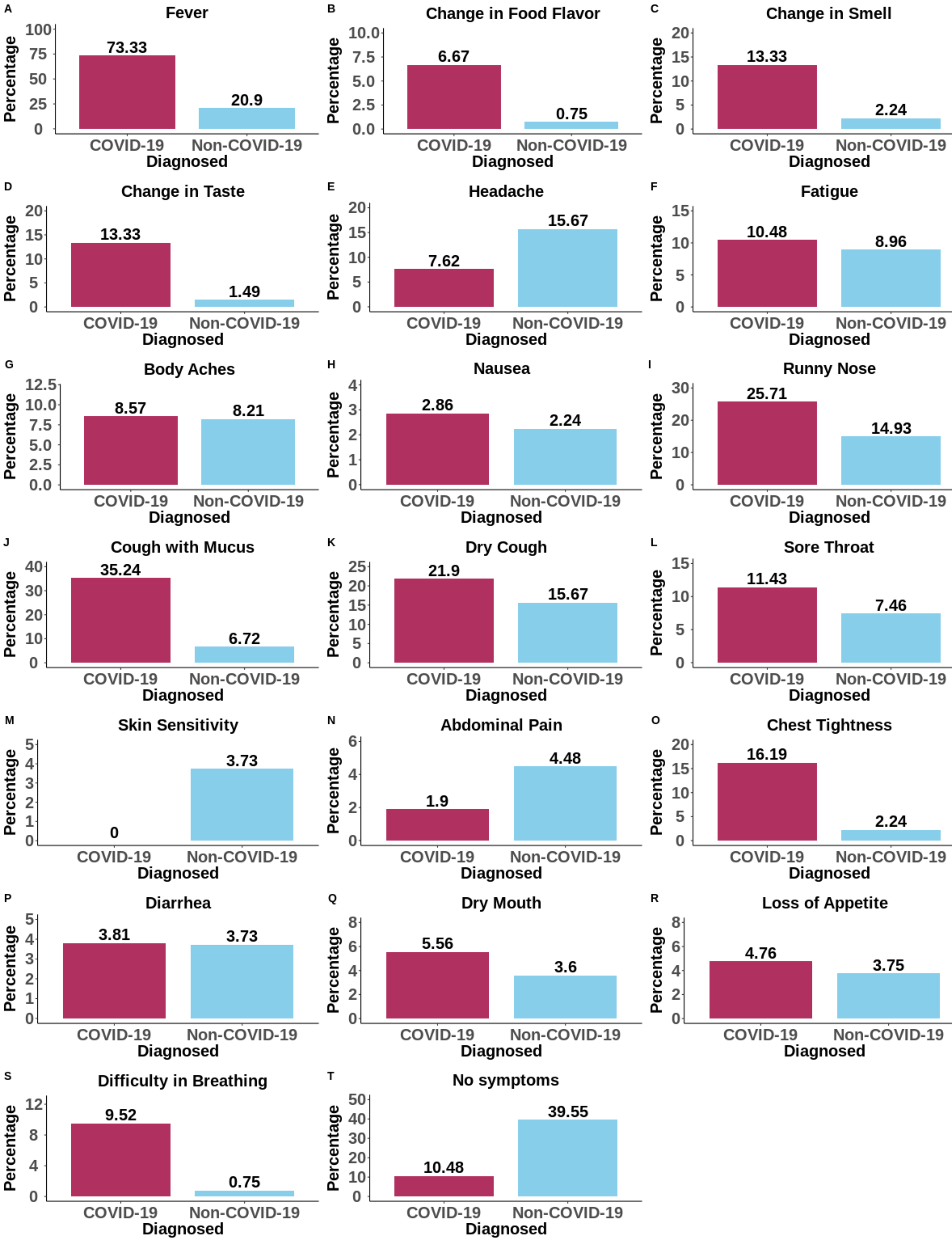


**Figure S4.** Reported symptoms of participants with and without COVID-19.

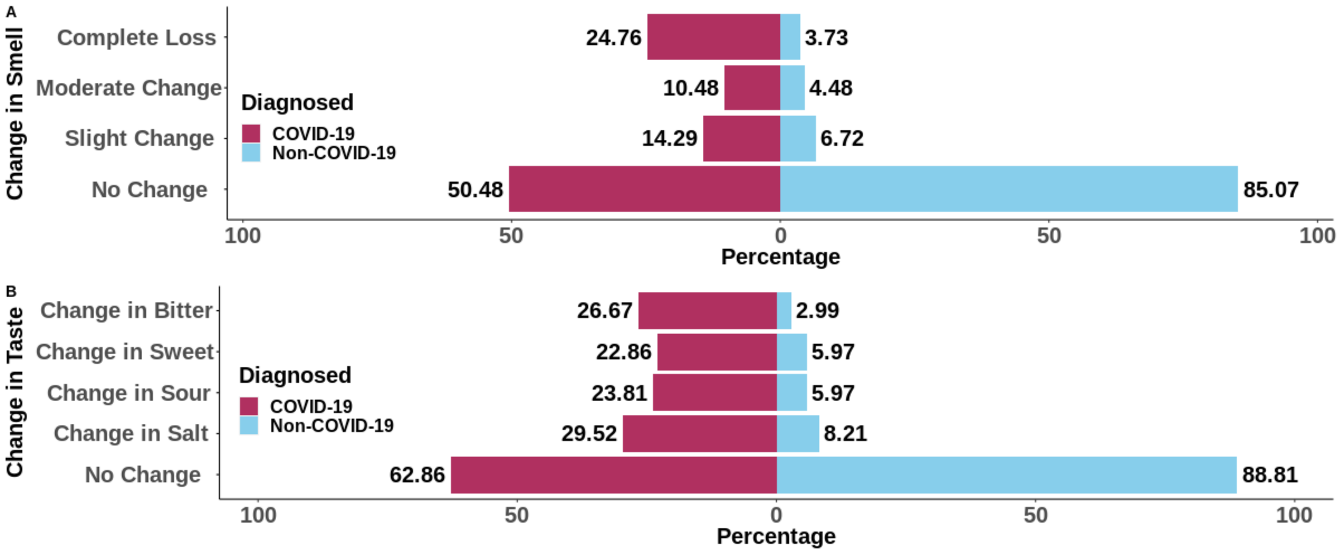


**Figure S5.** (A) Smell and (B) Taste specific symptoms in participants with and without COVID-19.
